# Wireless physical activity monitor use among adults living with HIV in a community-based exercise intervention study: a quantitative longitudinal observational study

**DOI:** 10.1101/2022.09.25.22280346

**Authors:** Joshua R. Turner, Justin Cheng, Judy Chow, Farhanna Hassanali, Hayley Sevigny, Michael Sperduti, Soo Chan Carusone, Matthieu Dagenais, Kelly K. O’Brien

## Abstract

**Objectives:** Our aim was to examine Wireless Physical Activity Monitor (WPAM) use and its associations with contextual factors (age, highest education level, social support, mental health) among adults living with HIV engaged in a community-based exercise (CBE) intervention.

**Design:** Quantitative longitudinal observational study.

**Setting:** Toronto YMCA, Ontario, Canada.

**Participants:** Eighty adults living with HIV who initiated the CBE intervention.

**Interventions:** Participants received a WPAM to track physical activity during a 25-week CBE intervention involving thrice-weekly exercise, supervised weekly (Phase 1) and a 32-week follow-up involving independent thrice-weekly exercise (Phase 2).

**Outcome measures:** Uptake was measured as participants who consented to WPAM use at intervention initiation. Usage was defined as the median proportion of days participants had great than 0 steps out of the total number of days in the study. We measured contextual factors using a baseline demographic questionnaire (age, highest education level), and median scores from the Medical Outcomes Study-Social Support Scale and Patient Health Questionnaire (mental health), where higher scores indicated greater social support and mental health concerns, respectively. We calculated Spearman correlations between WPAM usage and contextual factors defined as weak (ρ≥0.2, moderate (ρ≥0.4), strong (ρ≥0.6), or very strong (ρ≥0.8).

**Results:** Seventy-six of 80 participants (95%) consented to WPAM use. In Phase 1, 66% of participants (n=76) used the WPAM at least one day. Median WPAM usage was 50% (25^th^, 75^th^ percentile: 0%, 87%; n=76) of days enrolled in Phase 1 and 23% (0%, 76%; n=64) of days during Phase 2. Correlation coefficients ranged from weak for age (ρ=0.26) and mental health scores (ρ=-0.25) to no correlation (highest education level, social support).

**Conclusions:** Most adults living with HIV consented to WPAM use, however, usage declined over time. Future implementation of WPAMs should consider factors to promote sustained usage by adults living with HIV.

**Trial Registration:** NCT02794415

**STRENGTHS & LIMITATIONS OF THIS STUDY:**

- To our knowledge, this is the first quantitative study to measure wireless physical activity monitor (WPAM) use among adults living with HIV engaged in a community-based exercise intervention.
- The longitudinal study design enabled us to examine changes in WPAM use over time.
- Utilizing objective measures of physical activity (WPAM) and self-reported (questionnaire) measures of physical activity enabled us to investigate associations of different measurement approaches of physical activity levels among adults living with HIV.
- Limitations included variable and incomplete participant data across multiple data sources such as, WPAM synchronization, self-reported step count, and completion of weekly exercise questionnaires.

## INTRODUCTION

As the life expectancy of people living with HIV (PLWH) increases, individuals may experience more physical, mental and social health-related challenges attributed to HIV, multimorbidity, and aging.[1,2] These health-related challenges, known as disability, were described by PLWH as episodic and multidimensional, including physical, cognitive, emotional and social health domains.[3] Physical activity is an effective self-management strategy that can decrease risk of multimorbidity, and improve functional capacity, psychological well-being, and quality of life among PLWH.[4–6] Despite these benefits, variability exists in physical activity levels among PLWH.[7–9] This is consistent with the episodic nature of disability in PLWH and highlights the complexity of generalizing overall physical activity levels across this population. Variability in physical activity levels among PLWH may be due to barriers including HIV-associated stigma, stress, fatigue, low social support, and physical and mental health.[7,10] Community-based exercise (CBE) programs are one rehabilitation strategy to promote physical activity among PLWH. CBE programs are supervised, structured, and consist of individuals with similar characteristics (e.g., age, chronic conditions).[11] PLWH who participated in a supervised 6-month CBE program demonstrated improvements in cardiorespiratory fitness, strength, and flexibility.[12] However, PLWH have a greater dropout rate from physical activity interventions than other populations with chronic conditions.[13] Due to barriers to exercise faced by PLWH, technological devices could be incorporated into exercise interventions as an additional tool to measure and encourage consistent physical activity among PLWH.

Wireless physical activity monitors (WPAMs) are increasingly popular to promote and track physical activity.[14,15] WPAMs including commercially available pedometers and accelerometers (e.g., Fitbit™, Garmin, Apple Watch) can objectively measure and encourage physical activity in general and chronic illness populations.[14,16–18] WPAMs can track physical activity through measures of steps, distance, sedentary time, and energy expenditure.[18] Furthermore, WPAMs can facilitate self-monitoring and goal setting, which can motivate and reinforce positive behaviour change for physical activity participation.[16,19] However, the use of WPAMs may be limited by factors such as technological design or inability to capture features of physical activity (e.g., type or intensity of exercise, perceived exertion).[20]

The utility of WPAMs depends on the extent of their uptake and usage over time. Research on long-term use of WPAMs suggested that usage among adults in the general population declined over time.[21] Personal, psychosocial, environmental and technology related factors have been shown to influence WPAM uptake and usage in older adults.[22] Additionally, studies in the general population demonstrated that older age, and higher self-perceived health, education, and sense of community were positively correlated to adherence to WPAM use.[23,24]

WPAMs have been used to describe physical activity (e.g., intensity, heart rate, energy expenditure, distance walked) and pedometers to measure steps taken and distance walked in PLWH.[17] However, less is known regarding the extent of WPAM use and their potential to promote physical activity in this population.[17] Additionally, PLWH have a higher prevalence of lower levels of education[25] and social support,[10] and higher risk of mental health conditions[26] than the general population. Hence, the way in which these contextual factors may influence WPAM uptake and usage over time in PLWH, are unknown.

Our aim was to examine WPAM use among adults living with HIV engaged in a CBE intervention. Primary objectives were to 1) examine the extent of WPAM use among adults living with HIV engaged in a community-based exercise study, and to 2) examine the associations between WPAM usage and contextual factors including age, highest level of education, social support, and mental health among adults living with HIV. Secondary objectives were to 3) describe physical activity levels as measured by a WPAM and a self-reported physical activity questionnaire, and 4) explore the associations between self-reported (questionnaire) and objective measures (WPAM) of physical activity among adults living with HIV.

We hypothesized that older age, higher level of education, higher social support, and better self- perceived mental health would each demonstrate a positive moderate association (0.4 ≤ ρ ≤ 0.59)[27] with greater WPAM usage among adults living with HIV during a 25-week CBE intervention (objective 2). We hypothesized that self-reported physical activity would demonstrate a positive moderate association (0.4 ≤ ρ ≤ 0.59)[27] with greater WPAM-measured median daily step count, among adults living with HIV engaged in a 25-week CBE intervention (objective 4).

## METHODS

We conducted a quantitative longitudinal observational study using data from a community-based exercise (CBE) intervention study with adults living with HIV in Toronto, Canada. The CBE study included three phases: a 32-week baseline monitoring phase (Phase 0); a 25-week CBE intervention (Phase 1); and a 32-week follow-up monitoring period (Phase 2).[28] The focus of our study was on Phases 1 and 2 of the larger CBE study (Figure 1). Details of the CBE study protocol have been published elsewhere.[28]

**Figure 1.**
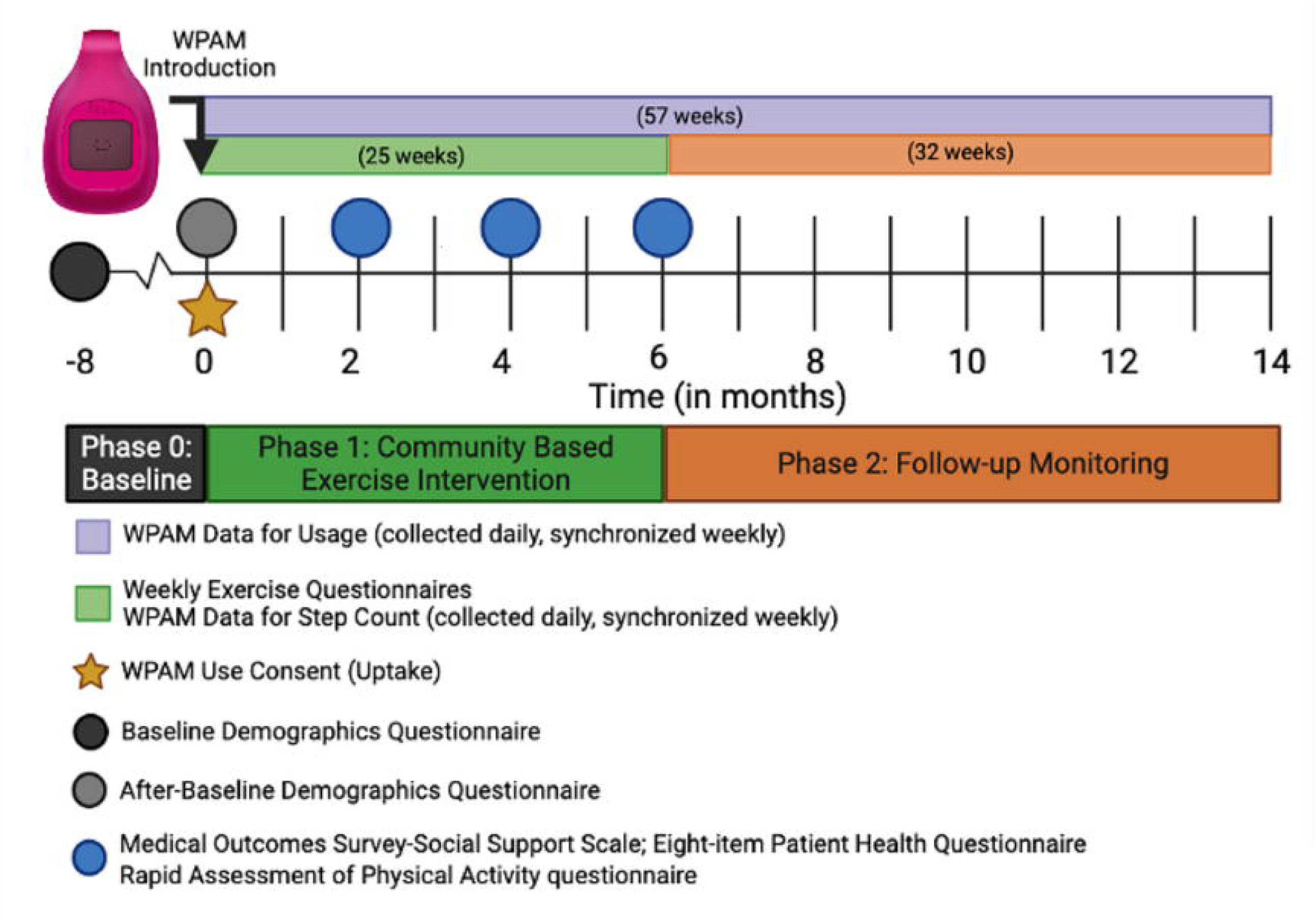
Study Timeline. Timeline of data sources collected during the Community-Based Exercise Study during Phase 0 (baseline, in black), Phase 1 (intervention phase, in green) and Phase 2 (follow-up phase, in orange). The Baseline Demographics Questionnaire was administered at the start of Phase 0 and the After-Baseline Demographics Questionnaire was administered at the initiation of Phase 1. The Medical Outcomes Survey-Social Support Scale (MOS-SSS), Eight-item Patient Health Questionnaire (PHQ-8), and Rapid Assessment of Physical Activity (RAPA) questionnaire were administered at baseline (month 0) and bi-monthly during Phase 1. Weekly exercise questionnaires were administered weekly and daily wireless physical activity monitor (WPAM) data were synchronized weekly.

### Participants

Participants in the CBE study[28] were recruited from the YMCA, community-based organizations and a specialty hospital. Participants included adults living with HIV (≥18 years) who considered themselves medically stable and safe to exercise upon completion of the Physical Activity Readiness Questionnaire.[29] This study included participants who enrolled in the CBE study and initiated Phase 1. Figure 1 outlines a timeline of the study components.

### Participant and Public Involvement Statement

We consulted participants living with HIV in the CBE study when developing the protocol for this study. These discussions helped shape our understanding of how participants interacted with the WPAM during the CBE study, informed our operationalization of study variables on WPAM use, and guided our investigation into contextual factors which may impact WPAM use, in relation to the lived experiences of PLWH.

### Community-Based Exercise Intervention

In Phase 1 (25-week CBE intervention), participants were provided with a YMCA membership and expected to participate in 90-minute multi-modal exercise sessions consisting of aerobic, strength, balance and flexibility thrice weekly, supervised weekly by a fitness instructor. In Phase 2 (32-week follow-up monitoring), participants received an 8-month extension to their YMCA membership and encouraged to continue independent exercise thrice weekly. Further details on the intervention were published elsewhere.[28,30]

#### Wireless Physical Activity Monitors

Participants were offered a Fitbit Zip™ at Phase 1 initiation. The Fitbit Zip™ is a WPAM that objectively measures daily step count (number of steps/day), distance walked (km), and calories burned (calories),[31] and a valid measure of step count in PLWH.[32] Participants who consented to WPAM use were asked to wear it all day on their waist. An online Fitbit™ account was created for each participant and instructions related to Fitbit™ setup, use, and synchronization were provided. Technical support was provided regarding synchronization as needed. Fitbit Zips™ lost during the study were not replaced. At the end of the study, participants were able to keep their Fitbit Zip™. Participants who possessed their own WPAM had the option to wear their own device. The type of WPAM used was recorded and daily data from Fitbit Zips™ and personal WPAMs downloaded if available.

### Data Sources / Measurements

The primary data source for the study was the WPAM, used to measure daily step count. Participants were asked to sync their WPAM weekly. Participants were asked to complete an online weekly exercise questionnaire throughout both phases of the study, to capture the type of WPAM used, total weekly step count, and total weekly distance walked (km). Additional data from self-reported questionnaires included: demographic and HIV characteristics, the Medical Outcomes Study-Social Support Scale (MOS-SSS; social support), the eight-item Patient Health Questionnaire (PHQ-8; mental health), and the Rapid Assessment of Physical Activity aerobic component (RAPA-1; aerobic physical activity) (Figure 1).

#### Uptake and Usage of Wireless Physical Activity Monitors

WPAM uptake was defined as the number and proportion of participants (out of the total number of participants in the study) who consented to WPAM use and sharing of data at Phase 1 initiation. WPAM usage was defined as the proportion of days each participant wore the WPAM (days with >0 steps measured) out of the total number of days enrolled in the study during Phase 1 (25 weeks), Phase 2 (32 weeks) and both phases combined (57 weeks). Reasons for WPAM non-usage were recorded in a participant spreadsheet during both phases.

#### Contextual Factors and Wireless Physical Activity Monitor Use

We examined associations between WPAM usage and age, highest level of education, social support, and mental health during Phase 1. We analyzed the associations during Phase 1 to maximize sample size. Participants’ age and highest level of education were reported in the baseline demographics questionnaire at CBE study initiation. Social support, defined as connectedness to one or multiple people who offer physical, emotional and relational support,[33] was measured by participants’ median MOS-SSS scores (range 19-95), with higher scores indicating greater perceived social support. Mental health, defined as a person’s emotional, psychological and social well-being,[34] was measured by participants’ median PHQ-8 scores (range 0-24), with higher scores indicating higher levels of mental distress. The MOS-SSS and the PHQ-8 have been previously used with PLWH.[26,35–37] The MOS-SSS and PHQ-8 were administered bi-monthly during Phase 1 (Figure 1).

#### Physical Activity

We measured aerobic physical activity objectively as participants’ median daily step count recorded by the WPAM in Phase 1 and by self-report as participants’ median RAPA-1 score during Phase 1. RAPA-1 scores range from 1-5, with higher scores indicating greater levels of activity. The RAPA-1 has demonstrated construct validity and responsiveness in adults living with HIV.[30] In addition, participants reported the weekly step count (number of steps/week) and distance walked (km) recorded by their WPAM on the weekly online exercise questionnaire throughout Phase 1.

#### Participant Characteristics

We measured participant characteristics including age, gender, living situation, source of income, and employment status using the baseline demographics questionnaire (Phase 0 initiation). We measured the number of concurrent health conditions, general health status, and exercise history from the demographic questionnaire completed at Phase 1 initiation (Figure 1).

### Analysis

We calculated the median (25^th^, 75^th^ percentile) for continuous variables (e.g., age, number of concurrent health conditions), and frequencies and percentages for categorical variables (e.g., gender, living situation, source of income, employment status, highest level of education, general health status, exercise history). We assessed the distribution of continuous and categorical study variables using the Shapiro-Wilk test, where α=0.05. For our correlation analyses, we interpreted correlation coefficients as very weak (ρ≤0.19), weak (0.2≤ρ≤0.39), moderate (0.4≤ρ≤0.59), strong (0.6≤ρ≤0.79), or very strong (ρ≥0.8)[27] with significance of α=0.05.

#### Wireless Physical Activity Monitor Uptake and Usage

We calculated the number of participants who consented to WPAM use, out of the total number of participants enrolled in Phase 1 (Uptake). We calculated the number of days each participant wore the WPAM out of the total number of days enrolled in the study, in Phase 1, Phase 2, and both phases combined (Usage). Usage was expressed as the median (25^th^, 75^th^ percentile) of days across all participants in the study. We reported the primary reasons for no WPAM usage in each phase.

#### Associations between Contextual Factors and Wireless Physical Activity Monitor Usage

We assessed the associations between WPAM usage during Phase 1 and a) age, b) highest level of education, c) median MOS-SSS score, and d) median PHQ-8 score for i) all participants and then ii) for all participants with non-zero WPAM usage. We calculated Spearman coefficients (ρ) and 95% confidence intervals (95% CI) for each of the four variables for non-normally distributed data.

#### Physical Activity

We calculated the median daily step count, measured by the WPAM, for each participant in Phase 1 and then calculated the median (25^th^, 75^th^ percentile) of participants’ median daily step count in Phase 1 for all participants with non-zero WPAM usage. We calculated the median RAPA-1 score for each participant in Phase 1 and then calculated the median (25^th^, 75^th^ percentile) of participants’ median RAPA-1 scores for all participants who completed at least one RAPA-1 questionnaire during Phase 1.

#### Associations between Objective and Self-Reported Measures of Physical Activity

We examined the association between each participant’s median daily step count, as measured by the WPAM, and median RAPA-1 scores in Phase 1. We calculated Spearman coefficients (ρ) and 95% CIs for non-normally distributed data.

We accounted for missing data or participants who were lost to follow-up by performing calculations with all data that were available, resulting in a smaller sample size for some calculations, which were reported when applicable. We used Microsoft Excel[38] and SPSS[39] to facilitate the statistical analysis.

### Sample Size

Eighty adults living with HIV initiated Phase 1, of which 76 (95%) consented to WPAM use. Thus, our maximum sample size to assess the associations of WPAM usage with contextual factors was 76 participants. Using a significance of α=0.05 and β=0.1, our sample size allowed us to detect a moderate correlation of ρ≥0.4 or greater.[40]

## RESULTS

Eighty participants initiated Phase 1, of which, 67 (84%) completed Phase 1, and 52 (65%) completed Phase 2 of the study.

### Wireless Physical Activity Monitor Uptake

Of the 80 participants who initiated Phase 1, 76 (95%) consented to WPAM use to monitor their physical activity (uptake). Of the 76 participants who consented to WPAM use, 64 (84%) remained enrolled in the study at the end of Phase 1, and 50 of the 64 participants (78%) who started Phase 2, completed the study. Figure 2 describes the retention of participants, and WPAM usage and uptake during the study.

**Figure 2.**
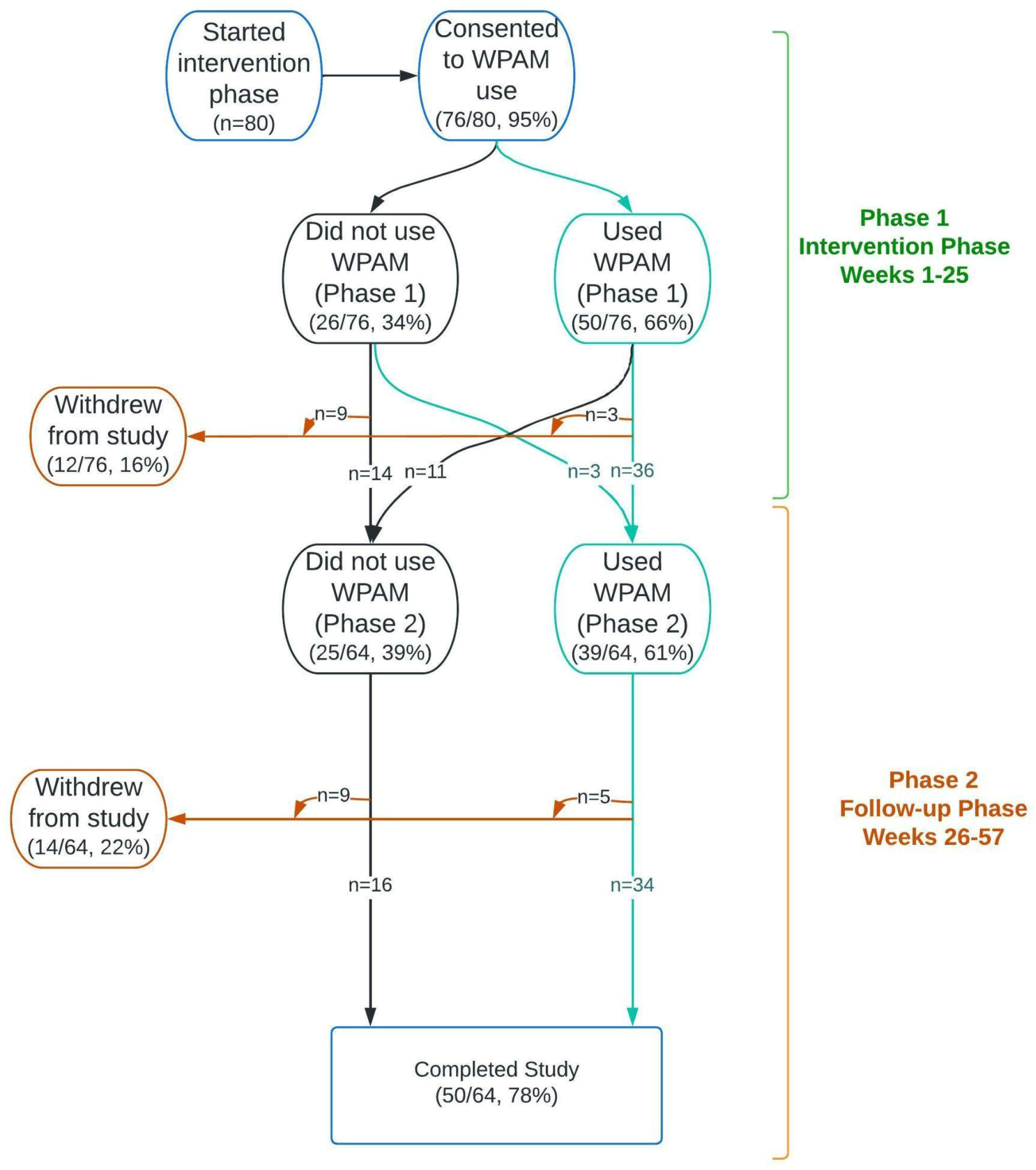
Participant Flow Chart. Flowchart of wireless physical activity monitor (WPAM) uptake (n=76), usage and retention of participants in Phase 1 (intervention phase) and Phase 2 (follow-up phase) of the Community-Based Exercise (CBE) Study.

### Participant Characteristics

Of the 76 participants who consented to WPAM use at Phase 1 initiation, the median age was 51 years (25^th^, 75^th^ percentile: 44, 59). The majority of the participants identified as a man (89%), white (68%), and lived alone (63%). Twenty-seven (36%) of participants were employed, while 36 (48%) reported their primary source of income was from financial assistance programs. The majority of participants (n=51; 53%) completed some form of post-secondary education or training. The median year of HIV diagnosis of the sample was 1999 (25^th^, 75^th^ percentile: 1989, 2008). Sixty-two participants (82%) reported living with two or more concurrent health conditions in addition to HIV, of which joint pain (46%) was the most common, followed by mental health concerns (39%) and muscle pain (39%). Thirty-six (47%) participants reported exercising 3 to 5 days per week prior to initiating Phase 1 of the CBE study. Participant characteristics for the 50 participants who had non-zero WPAM usage were similar to the characteristics of the entire sample described above (Table 1).

**Table 1.** Characteristics of participants who initiated the community-based exercise (CBE) study and agreed to use a Wireless Physical Activity Monitor (WPAM)

| Participant Characteristic | Participants who consented to WPAM use at the start of the intervention (n=76) | Participants who used a WPAM during the intervention (n=50) |
| --- | --- | --- |
| **PERSONAL CHARACTERISTICS** | **Median (25<sup>th</sup>, 75<sup>th</sup> percentile)** | **Median (25<sup>th</sup>, 75<sup>th</sup> percentile)** |
| **Age at study initiation (years)** | 51 (44, 59) | 53 (45, 60) |
|  | # (%) | # (%) |
| **Number of participants ≥50 years of age** | 44 (58%) | 32 (64%) |
| **Gender** | # (%) | # (%) |
| Woman | 6 (8%) | 5 (10%) |
| Man | 68 (89%) | 44 (88%) |
| Other (intersexed, non-binary) | 2 (3%) | 2 (2%) |
| **Race and/or Ethnicity** | # (%) | # (%) |
| White | 52 (68%) | 34 (68%) |
| Asian | 14 (18%) | 8 (16%) |
| Indigenous | 3 (4%) | 2 (4%) |
| Black or African | 5 (7%) | 4 (8%) |
| Hispanic | 3 (4%) | 3 (6%) |
| Other (Arab, Jewish, Mediterranean, Indo-Caribbean) | 5 (7%) | 3 (6%) |
| **Living situation** | # (%) | # (%) |
| Living alone | 58 (76%) | 37 (74%) |
| **Primary source of income at study initiation** | # (%) | # (%) |
| Employer | 22 (29%) | 14 (28%) |
| Pension | 17 (23%) | 10 (20%) |
| Financial Assistance (Ontario Disability Support Program, Employment Insurance and Ontario Works) | 36 (48%) | 26 (52%) |
| **Employment status at study initiation** | **# (%)** | **# (%)** |
| Employed | 27 (36%) | 16 (32%) |
| Retired | 7 (9%) | 5 (10%) |
| Unemployed | 42 (55%) | 29 (58%) |
| **Highest level of education completed at study initiation** | **# (%)** | **# (%)** |
| Less than high school diploma | 9 (12%) | 6 (12%) |
| Completed High School | 2 (3%) | 0 (0%) |
| Some trade/technical training, college or university | 14 (18%) | 12 (24%) |
| Trade/technical training, college or university completed | 35 (46%) | 20 (40%) |
| Completed post-graduate degree | 16 (21%) | 12 (24%) |
| **Self-reported general health status at CBE initiation** | **# (%)** | **# (%)** |
| Excellent | 4 (5%) | 1 (2%) |
| Very good | 23 (30%) | 18 (36%) |
| Good | 32 (42%) | 21 (42%) |
| Fair | 16 (21%) | 9 (18%) |
| Poor | 1 (1%) | 1 (2%) |
| **Smoking history at CBE Initiation** | **# (%)** | **# (%)** |
| Current Smoker (in past 30 days) | 12 (16%) | 7 (14%) |
| Former smoker | 34 (45%) | 24 (48%) |
| Never Smoked | 27 (36%) | 17 (34%) |
| Don't know or prefer not to answer | 3 (4%) | 2 (4%) |
| **Patient Reported Outcome Measures during CBE intervention** | **Median (25<sup>th</sup>, 75<sup>th</sup> percentile)** | **Median (25<sup>th</sup>, 75<sup>th</sup> percentile)** |
| Medical Outcome Survey-Social Support Scale Social Support score (range 19-95) (n=69, 49) | 58 (36, 76) | 56 (29, 72) |
| 8-Item Patient Health Questionnaire score (range 0-24) (n=69, 49) | 6 (3, 10) | 5 (2, 9) |
| **HIV CHARACTERISTICS** |  |  |
| **Year of HIV diagnosis** | **Median (25<sup>th</sup>, 75<sup>th</sup> percentile)** | **Median (25<sup>th</sup>, 75<sup>th</sup> percentile)** |
|  | 1999 (1989, 2008) | 1998 (1989, 2008) |
|  | # (%) | # (%) |
| **Viral Load Undetectable** (<50 copies/mL) at CBE initiation (n=75, 49) | 73 (97%) | 48 (98%) |
| **Receiving HIV Care** at CBE initiation | 75 (99%) | 49 (98%) |
| **Median total # of concurrent health conditions in addition to HIV** | **Median (25<sup>th</sup>, 75<sup>th</sup> percentile)** | **Median (25<sup>th</sup>, 75<sup>th</sup> percentile)** |
|  | 5 (2,7) | 4 (2, 7) |
| **Most commonly self-reported concurrent health conditions at CBE initiation (>30%)** | **# (%)** | **# (%)** |
| Joint pain (arthritis) | 35 (46%) | 24 (48%) |
| Mental health (e.g. depression, anxiety) | 30 (39%) | 20 (40%) |
| Muscle pain | 30 (39%) | 19 (38%) |
| Bone and joint disorder | 24 (32%) | 16 (32%) |
| **Living with ≥2 concurrent health conditions** | 62 (82%) | 42 (84%) |
| **EXERCISE CHARACTERISTICS** |  |  |
| **Days exercising per week at CBE Initiation** | **# (%)** | **# (%)** |
| 0 days | 13 (17%) | 5 (10%) |
| 1-2 days | 18 (24%) | 11 (22%) |
| 3-5 days | 36 (47%) | 26 (52%) |
| 6-7 days | 9 (12%) | 8 (16%) |
| **Patient Reported Outcome Measure of Physical Activity** | **Median (25<sup>th</sup>, 75<sup>th</sup> percentile)** | **Median (25<sup>th</sup>, 75<sup>th</sup> percentile)** |
| Rapid Assessment of Physical Activity aerobic component score (range 1-5) (n=69, 49) | 5 (4, 5) | 5 (5, 5) |

n=76 consented at start of intervention, and n=50 usage during the intervention for all characteristics, unless otherwise stated.

### Wireless Physical Activity Monitor Devices Used

Of the 76 participants who consented to WPAM use at Phase 1 initiation, 36 (47%) provided a description of their WPAM device in at least one weekly exercise questionnaire. Twenty-five (69%) of the 36 participants reported using the Fitbit Zip™ exclusively during Phase 1 and Phase 2, while 6 (8%) participants reported using a combination of the Fitbit Zip™ and another WPAM (generic step counter, unspecified Fitbit™ device, wristband pedometer, Samsung Gear, iPhone, Fitbit™ Charge 2). Five (14%) of the 36 participants reported they used a personal WPAM during both phases, which included an Apple Watch (n=1), Fitbit Flex 2™ (n=2), Fitbit Ionic™ (n=1), and an unspecified Fitbit™ device (n=1).

### Wireless Physical Activity Monitor Usage

Of the 76 participants who consented to WPAM use, 50 (66%) used their device at some point during Phase 1. Of the 64 participants remaining in Phase 2, 39 (61%) used their WPAM at some point during Phase 2. Three participants who never used the WPAM in Phase 1 began using it in Phase 2. The median WPAM usage during Phase 1 was 50% (25^th^, 75^th^ percentile: 0%, 87%; n=76) of days out of the total number of days enrolled in Phase 1, and 23% (0%, 76%; n=64) of days during Phase 2 (Figure 3). Median usage over the two phases combined was 31% (0%, 73%; n=76) of days out of the total number of days enrolled in Phase 1 and 2. When the participants with no WPAM usage were removed from analysis, the proportion of WPAM usage was 79% (51%, 93%, n=50) during Phase 1, 64% (31%, 87%, n=39) during Phase 2 and 54% (23%, 84%, n=53) when both phases were combined together. Participant’s WPAM usage declined over time (Supplemental File 1).

**Figure 3.**
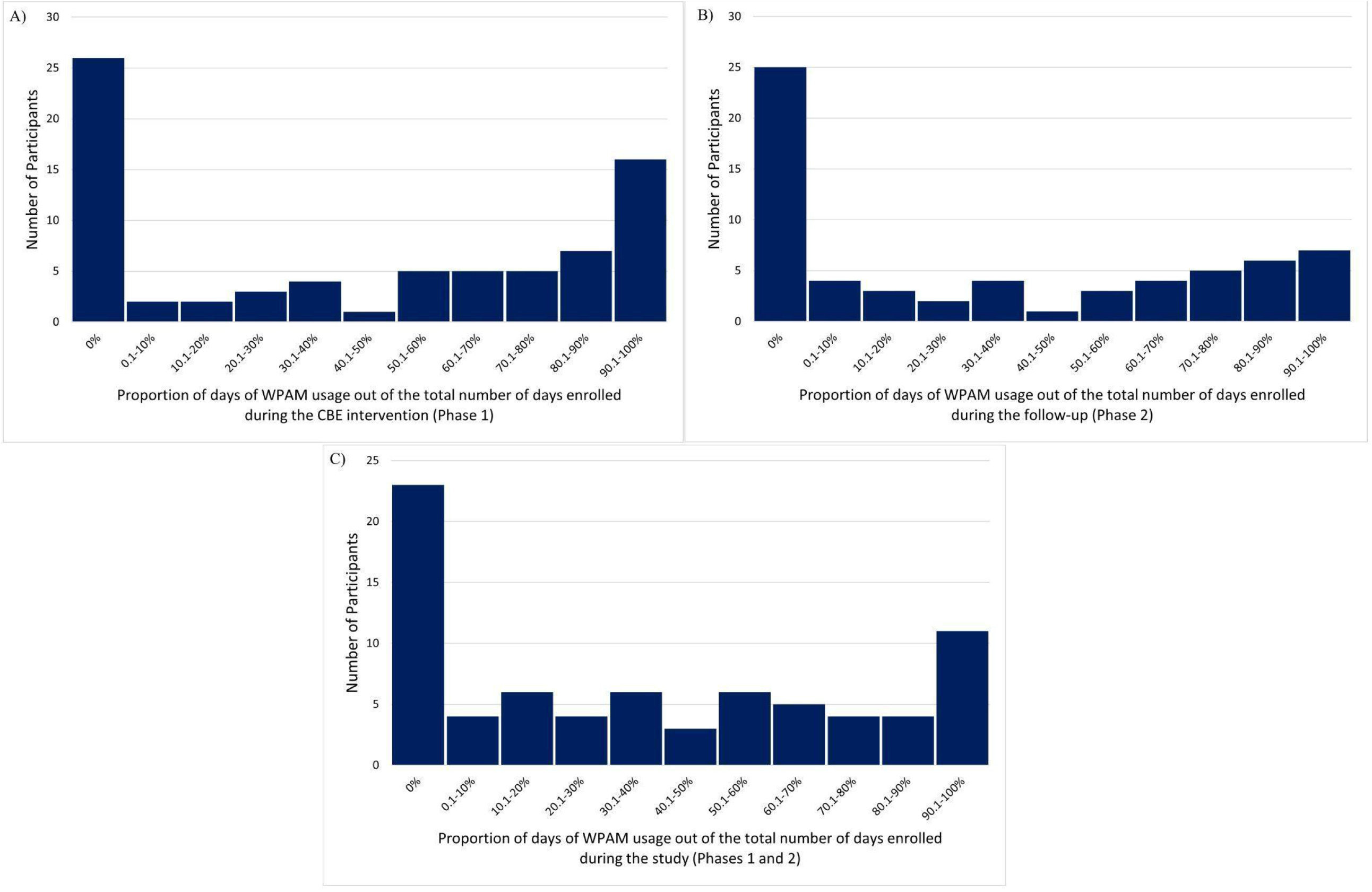
Histograms of Wireless Physical Activity Monitor Usage. Histograms of the proportion of days of Wireless Physical Activity Monitor (WPAM) usage out of the total number of days all participants (with or without WPAM usage) were enrolled during A) Phase 1 (intervention phase, n=76), B) Phase 2 (follow-up phase, n=64), and C) Phase 1 and 2 combined (n=76).

### Reasons for Wireless Physical Activity Monitor Non-usage

Twenty-six participants (34%) did not use the WPAM during Phase 1, and 25 participants (39%) did not use the WPAM in Phase 2. The most common reason reported for WPAM non-use in Phase 1 was account or login issues (42%) and previous discontinuation of WPAM use during the intervention (36%) in Phase 2 (Supplemental File 2).

### Associations of Contextual factors and Wireless Physical Activity Monitor Usage

Among participants who consented to WPAM use, age demonstrated a weak positive correlation with WPAM usage (ρ=0.26; p=0.02; 95% CI: 0.03 to 0.46; n=76). Self-reported mental health (median PHQ-8 scores) demonstrated a weak, negative correlation with WPAM usage (ρ=-0.25; p=0.04; 95% CI: -0.47 to -0.01) meaning higher levels of mental health concerns were associated with less WPAM usage. WPAM usage was not associated with highest level of education (ρ=0.06; p=0.63; 95% CI: -0.18 to 0.29) or social support (median MOS-SSS scores) (ρ= -0.17; p=0.16; 95% CI: -0.40 to 0.07) (Figure 4). When participants with no WPAM usage were removed from analysis, the results were similar (Supplemental File 3).

**Figure 4.**
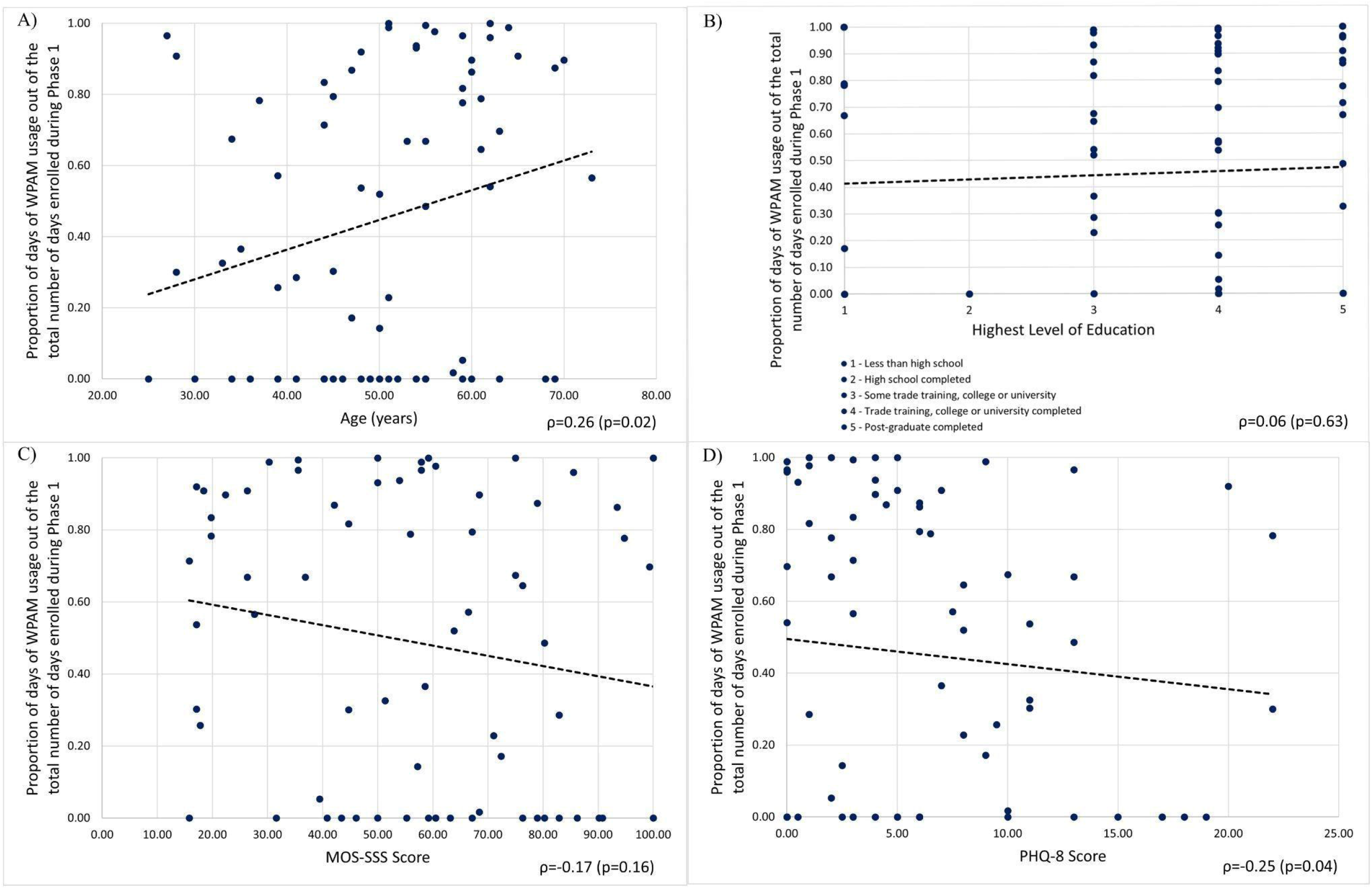
Scatter Plots of Wireless Physical Activity Monitor Usage and Contextual Factors Associations. Correlations between the proportion of days of wireless physical activity monitor (WPAM) usage out of the total number of days enrolled in Phase 1 (intervention phase) of all participants (with and without WPAM usage) and A) age (n=76), B) highest level of education (n=76), C) social support as assessed by the Medical Outcomes Survey-Social Support Scale (MOS-SSS) (n=69), and D) mental health concerns as assessed by the eight-item Patient Health Questionnaire (PHQ-8) (n=69).

### Physical Activity

During Phase 1, 50 of the 76 participants (65%) had greater than 0% WPAM usage, of which the median of participants’ median daily step count was 9,540 steps (25^th^, 75^th^ percentile: 6,121, 10,990). During Phase 1, 69 participants completed at least one RAPA-1 questionnaire, of which the median of participants’ median RAPA-1 value was 5 (25^th^, 75^th^ percentile: 4, 5) out of 5 indicating an “active’’ physical activity status, exercising at moderate to vigorous intensity. Of the 76 participants consenting to WPAM use, 15 participants self-reported their weekly step count and 16 self-reported their weekly distance walked during Phase 1 in a weekly exercise questionnaire. The median of participants’ median weekly step count was 57,729 steps (25^th^, 75^th^ percentile: 1,655, 70,684) and the median of participants’ median weekly distance walked was 39km (25^th^, 75^th^ percentile: 4, 49).

### Associations of Self-Report and Objective Measures of Physical Activity

Participants’ median daily step count and RAPA-1 scores were non-normally distributed. Hence, we calculated a Spearman correlation to assess the association between participants’ objective measure of physical activity (median daily step count as measured by the WPAM) with participants’ self-reported physical activity (median RAPA-1 scores). Forty-nine participants during Phase 1, had step counts recorded by their WPAM and completed the RAPA-1 at least once. Median daily step count as measured by the WPAM was not associated with RAPA-1 scores (ρ=0.14; p=0.33; n=49; 95% CI: -0.15 to 0.41).

## DISCUSSION

Although WPAMs have been used in previous literature to quantify physical activity in PLWH, to our knowledge, this is the first study to examine the extent of WPAM uptake and usage in PLWH. We found WPAM uptake was high where the majority of participants (95%, n=76) consented to WPAM use. This is consistent with a previous study by Hassani et al. (2014) that investigated consent to WPAM use for a 9-day period in older adults.[41] Participants were provided with a WPAM as part of the intervention and researchers found the rate of consent was 92%.[41] As uptake in our study and Hassani et al. (2014) was measured in the context of a research study where participants were provided with a WPAM, this may have influenced participants’ willingness to use. Therefore, it is important to consider how to promote their use in a real world setting. When different WPAMs were compared among older adults, aesthetics, price, and comfort were factors that positively impacted the acceptability of devices.[42] Due to HIV-related stigma that can be experienced with HIV disclosure among PLWH, privacy concerns may be a prevalent barrier to WPAM uptake as devices are often synced to a website or mobile app. In a survey that investigated the use of health apps and smart devices (including WPAMs) in PLWH, only 22% of participants owned a smart device.[43] The main concern surrounding the use of technology was privacy of personal information.[43] Thus, further research on factors that facilitate or hinder WPAM uptake in PLWH is necessary to understand how they can be encouraged among this population.

WPAM usage in this study was variable across the population despite high uptake (Figure 3). Median usage decreased from 50% in Phase 1 to 23% in Phase 2. Our findings are consistent with Hermsen et al. who found WPAM usage in the general population declined over a period of 320 days with approximately 50% of participants still using the WPAM after 6 months.[21] Decline in usage over time may be attributed to decreased novelty of the WPAM, which was associated with a decrease in interest or competing life demands.[44]. External reminders and feedback related to goal setting via text messaging may be a strategy to combat the decline in usage as seen in a study that investigated the experiences of WPAM usage in older adults.[19] Additionally, factors associated with long-term WPAM use beyond the novelty stage includes existing personal motivators (e.g. goals to be physically active to improve health) and social support.[44] Despite the potential novelty of WPAMs wearing off, a subset of our participants used the WPAM frequently throughout the study (Figure 3C). This demonstrates that sustained WPAM use is possible in PLWH. However, solely providing participants with WPAMs may not be sufficient to support sustained use and may be supplemented with additional strategies over time.

Although technical support was provided to participants in our study, the most prevalent reason for WPAM non-usage were account or login issues (Supplemental File 2). WPAM usage was likely underestimated in our study as participants may have used their WPAMs without synchronizing it. These results highlight the potential difficulties around technology literacy and comfort in using mobile apps or computers to synchronize WPAMs, particularly in older adults.[20] This may deter their use outside of a research context, as individuals may need to seek technical support from providers independently, or from community and social support networks.[45] Thus, the availability of reliable technical support and device education may help to maintain sustained WPAM use among PLWH.

Our results found weak to no associations between WPAM usage and age, highest level of education, mental health, and social support scores. While the coefficients for age and mental health were statistically significant, the confidence intervals of the estimates approached zero, indicating no association. In the general population, older age, better self-perceived health and higher levels of education have been positively correlated with increased accelerometer use.[24] In contrast, economically disadvantaged persons with lower physical and mental health and sense of community were correlated with less WPAM use.[23] These studies, however, investigated the effects of these contextual factors in conjunction as a predictor of WPAM use, whereas our study investigated the factors independently. Experiences of PLWH and the resulting disability are multidimensional including physical, social and emotional domains.[46] Personal and environmental contextual factors such as social support, stigma, and personal attributes (e.g., age, comorbid illness) all interact to influence disability among this population.[46] These intersections may influence one another to impact WPAM usage. Li et al. suggested personal, psychosocial, environmental and technology-related factors collectively contributed to the long-term use of WPAMs in older adults.[22] Intrinsic motivation was reported as the most prevalent factor for use, however, external support from friends and family and feedback on goals from the device also supported their usage.[22] Thus, researchers and clinicians should consider the collective experiences of PLWH, the various factors which may influence this, and how these factors may interact with each other when choosing and implementing WPAMs.

This study focused on a small subset of contextual factors which may influence usage in PLWH while other factors should also be considered. As mentioned, HIV-related stigma may influence participants’ usage of technology due to concerns of privacy,[43] therefore strategies to address these barriers may support usage. Also, PLWH have previously described their experiences of disability as episodic, with unpredictable periods of illness and wellness.[46] Investigations into WPAM usage corresponding to periods of episodic disability may speak to the usability of WPAM in PLWH over sustained periods of time.

### Self-Reported and Objective Physical Activity

The median of the participants’ median daily step count was 9,540 steps across the 25-week intervention. This level of physical activity was similarly reported by Cook et al. where 60% of PLWH performed greater than 10,000 steps/day as measured by a WPAM.[7] In a different study with older adults living with HIV, the average daily step count was 3,543 when not engaged in an exercise intervention and increased by 43% during the intervention.[47] This demonstrates that step-based activities can be an accessible mode of aerobic exercise for PLWH.[7,48] The recommended daily step count for older, chronically ill adults is approximately 7,000-10,000 steps/day[49] and a moderate pace is the minimum intensity required to gain cardiovascular benefits.[50] Although the median step count of the participants in this study was within the recommended range,[49] the Fitbit Zip™ was unable to capture the intensity of physical activity. Intensity is important as the Canadian Physical Activity Guidelines recommends adults to engage in at least 150 minutes of moderate to vigorous aerobic physical activity per week.[50] Quigley et al. are currently investigating healthy lifestyle living and goal management training among PLWH[51] using a WPAM (Garmin VivoFit 4) to record aerobic intensity to determine achievement of physical activity guidelines.[51] Therefore, WPAMs that accurately measure step count and aerobic intensity may help researchers and clinicians in determining if PLWH are meeting physical activity guidelines.

When investigating a participant’s self-reported physical activity, the median of the participants’ median self-reported RAPA-1 (aerobic) scores was 5, indicating that participants commonly reported themselves as “active’’, exercising at moderate to vigorous intensity. However, we found no correlation between the WPAM-measured daily step count and the RAPA-1 scores. The absence of a correlation may be attributed to a ceiling effect of the RAPA-1 scores among the sample as they were engaged in a structured exercise intervention. Nevertheless, self-reported measures of physical activity can provide information including perceived exertion, upper extremity movement, and swimming measures, which may not be obtained by WPAMs.[52] Validated self-reported measures used in conjunction with WPAMs may provide supplementary qualitative information related to physical activity. A combination of measures could equip researchers and clinicians with a comprehensive toolset to aid physical activity prescription, progression, and monitoring among PLWH.

### Limitations

Our study is not without limitations. Uptake was likely overestimated as participants were provided a Fitbit Zip™ as part of the intervention. Additionally, the sample was predominantly white men, 50 years or older, living in Toronto, Canada. Therefore, the transferability of findings to the broader HIV population including women, PLWH in rural or remote areas without WiFi access, or those with financial constraints, is unknown. Furthermore, our fixed sample size of 76 limited our ability to detect correlations less than 0.4.[40] While our analysis was descriptive and exploratory, results contribute to a broader understanding of WPAM use and may inform their implementation in research and clinical interventions. Another limitation was missing data across the data sources (self-report and WPAM) and across time points in the study. Strategies used to maximize available data included calculating median scores of the MOS-SSS, PHQ-8, and RAPA-1 questionnaires during Phase 1, and including participants with any WPAM data available irrespective of the type of WPAM used. Finally, the WPAM used in this study, the Fitbit Zip™, is clipped onto the hip and is not waterproof, limiting its ability to measure upper extremity activities as opposed to other WPAMs worn on the wrist.[31] While the Fitbit Zip™ is no longer available for commercial purchase, the variable reasons for WPAM non-use discussed in our study are not exclusive to the Fitbit Zip™, and may be used to inform strategies to promote sustained use of any commercially available WPAM.

## CONCLUSIONS

While the majority of adults living with HIV consented to WPAM use, WPAM usage was variable and declined over time. Furthermore, WPAM usage was not associated with age, highest level of education, mental health or social support and a self-reported measure of aerobic physical activity was not associated with WPAM-measured step count. Consequently, we recommend that future implementation of WPAMs in research or clinical settings should consider factors which may promote sustained WPAM usage, including providing technical support and incorporating goal setting. While the contextual factors in our study did not appear to impact WPAM usage in isolation, future research should investigate how contextual factors may interact to affect WPAM usage. Finally, combining self-reported physical activity measures with WPAMs may provide a more complete understanding of physical activity among PLWH.

## Data Availability

All data produced in the present work are contained in the manuscript.

## ACKNOWLEDGEMENTS

This research was completed in partial fulfillment of the requirements for an MScPT degree at the University of Toronto. The authors thank the adults living with HIV who participated in the Community-Based Exercise (CBE) study. The authors acknowledge members of the original CBE study research team involved in study design, funding acquisition, data collection, and analysis of primary outcomes (Aileen M. Davis, Ada Tang, Patricia Solomon, Ahmed M. Bayoumi, Lisa Avery) and representatives from the YMCA who facilitated the CBE study implementation (Mehdi Zobeiry, Zoran Pandovski, Ivan Illic). We also acknowledge two community partner experts familiar with the CBE study, Greg Robinson and Shaz Islam, who met with us to enhance our understanding of the larger CBE study and specific considerations of the Fitbit Zip™ in the context of our study.

## FOOTNOTES

JRT, JC, JC, FH, HS, and MS all contributed equally to the study.

## AUTHORS’ CONTRIBUTIONS

KKO, and SCC were involved in the original CBE study team involved in the conceptualization of the study design and data collection. KKO, SSC, and MD provided guidance throughout all stages of the research including development of the research protocol, analysis, interpretation of results, and manuscript development. MD provided assistance with the SPSS statistical analyses. KKO, SCC, and MD supervised JRT, JC, JC, FH, HS, and MS who developed the protocol, analyzed the data, and drafted the manuscript in partial fulfilment of requirements for an MScPT degree at the University of Toronto. JRT, JC, JC, FH, HS, and MS (MScPT students) developed skills in quantitative research methodology in course work on ethics, research strategies, literature review, study design, protocol development, and learning to use SPSS statistical software. All steps were closely reviewed and guided by advisors (KKO, SCC, MD). KKO is the senior responsible author and guarantor of the study. All authors read and approved the final manuscript.

## PARTICIPANT CONSENT

All participants in this study provided written informed consent to participate in the CBE study and to WPAM use.

## ETHICS APPROVAL

This study was approved by the University of Toronto Health Sciences Research Ethics Board (Protocol #41886; Supplemental File 4). All participants provided written informed consent to participate in the CBE study and subsequently asked to provide written consent to WPAM use (Supplemental File 5).

## DATA AVAILABILITY STATEMENT

The data supporting the conclusions of this article are included within the article and its supplemental files. The data used and/or analysed during the current study are available from the corresponding author on reasonable request.

## SUPPLEMENTAL FILES

Supplemental File 1 – Weekly Wireless Physical Activity Monitor Usage

Supplemental File 2 – Reasons for Non-usage of Wireless Physical Activity Monitors

Supplemental File 3 – Scatter Plots of Wireless Physical Activity Monitor Usage and Contextual Factors Associations with Zero-Usage Participants Removed

Supplemental File 4 – Research Ethics Board Study Protocol and Approval Letter

Supplemental File 5– Community Based Exercise Study Participant Consent Form

