## Supplemental File 1 for "Wireless physical activity monitor use among adults living with HIV in a community-based exercise intervention study: a quantitative longitudinal observational study"

Supplemental File 1 - Wireless physical activity monitor use among adults living with HIV in a community-based exercise intervention study: a quantitative longitudinal observational study

**Supplemental File 1 - Weekly Wireless Physical Activity Monitor Usage**

Number of participants who recorded at least one day of Wireless Physical Activity Monitor (WPAM) usage (navy blue bar) during each week of the study. The height of the gray bar represents the total number of participants enrolled in the study each week.

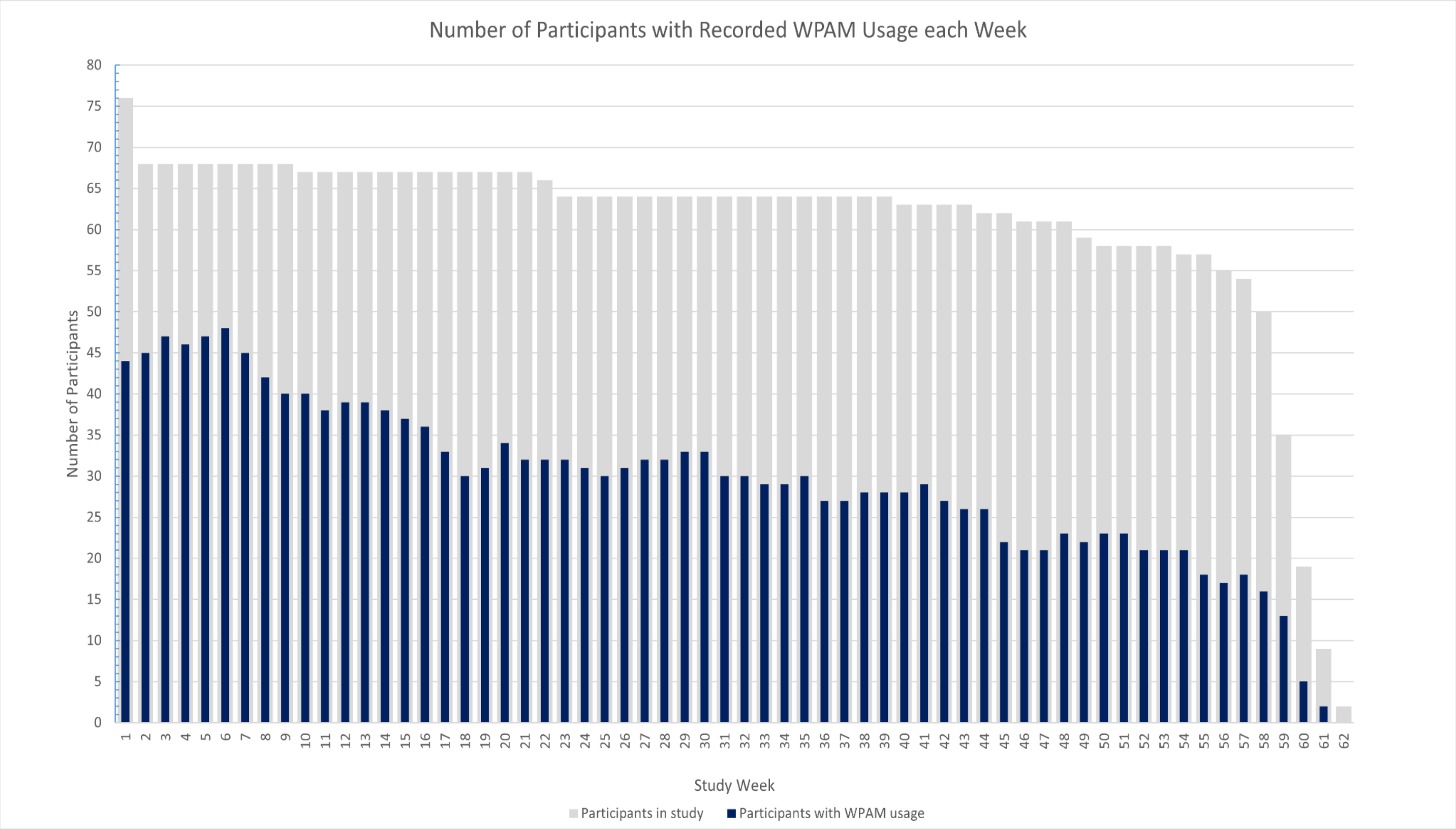


Date Last Revised: September 25, 2022
