## Supplemental File 2 for "Wireless physical activity monitor use among adults living with HIV in a community-based exercise intervention study: a quantitative longitudinal observational study"

Supplemental File 2 - Wireless physical activity monitor use among adults living with HIV in a community-based exercise intervention study: a quantitative longitudinal observational study

**Supplemental File 2 - Reasons for Non-usage of Wireless Physical Activity Monitors**

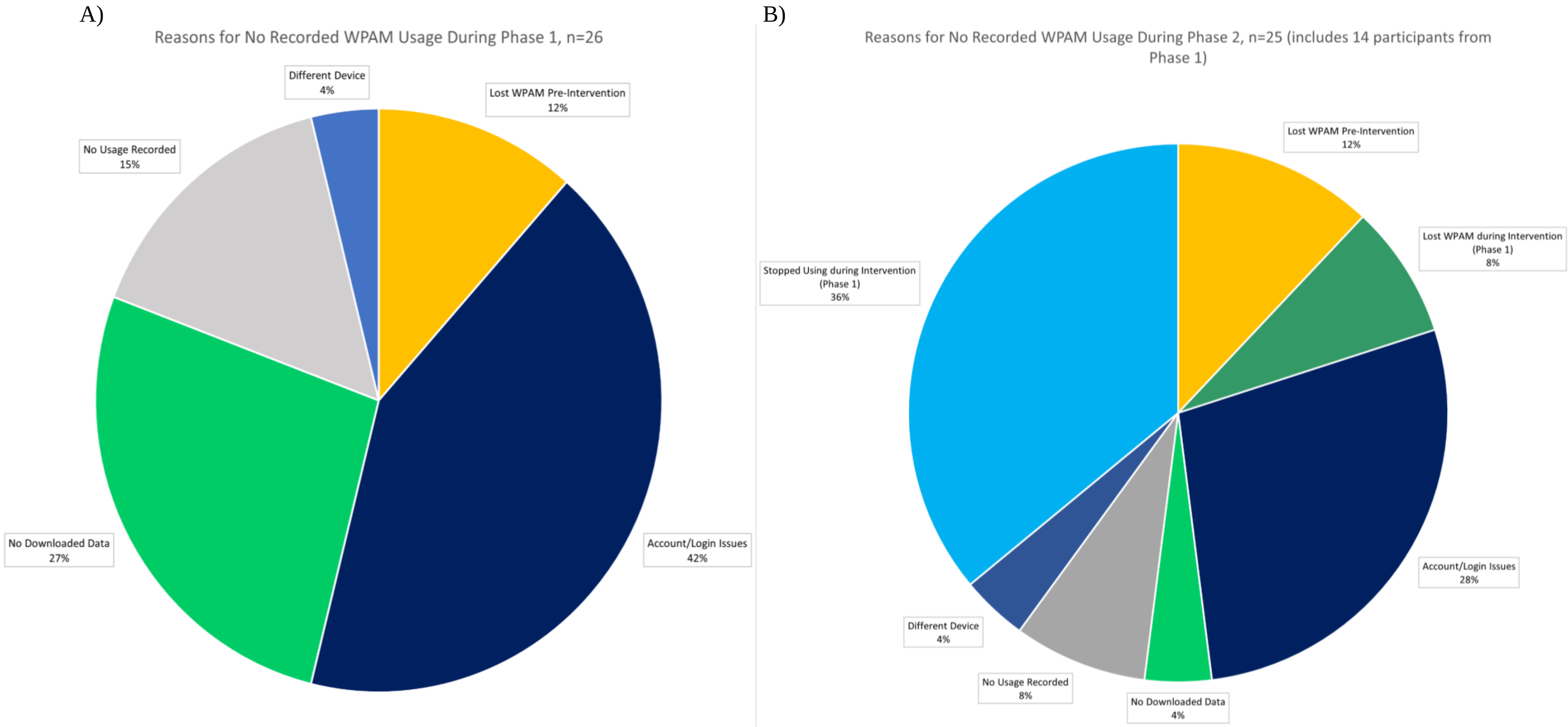


**Supplemental File 2 - Reasons for Non-usage of Wireless Physical Activity Monitors**

The reasons that participants had no recorded WPAM usage during A) the intervention phase (Phase 1) (n=26), and B) the follow-up phase (Phase 2) (n=25) of the study. Fourteen participants included in Figure A are also included in Figure B since these participants had no recorded WPAM usage during both phases.

Date Last Revised: September 25, 2022
