## Supplementary material for "Wireless physical activity monitor use among adults living with HIV in a community-based exercise intervention study: a quantitative longitudinal observational study": Supplemnetal File 3

Supplemental File 3 - Wireless physical activity monitor use among adults living with HIV in a community-based exercise intervention study: a quantitative longitudinal observational study

**Supplemental File 3 - Scatter Plots of Wireless Physical Activity Monitor Usage and Contextual Factors Associations with Zero-Usage Participants Removed**
Correlations between the proportion of days of wireless physical activity monitor (WPAM) usage out of the total number of days enrolled and contextual factors (age, highest level of education, social support and mental health scores).

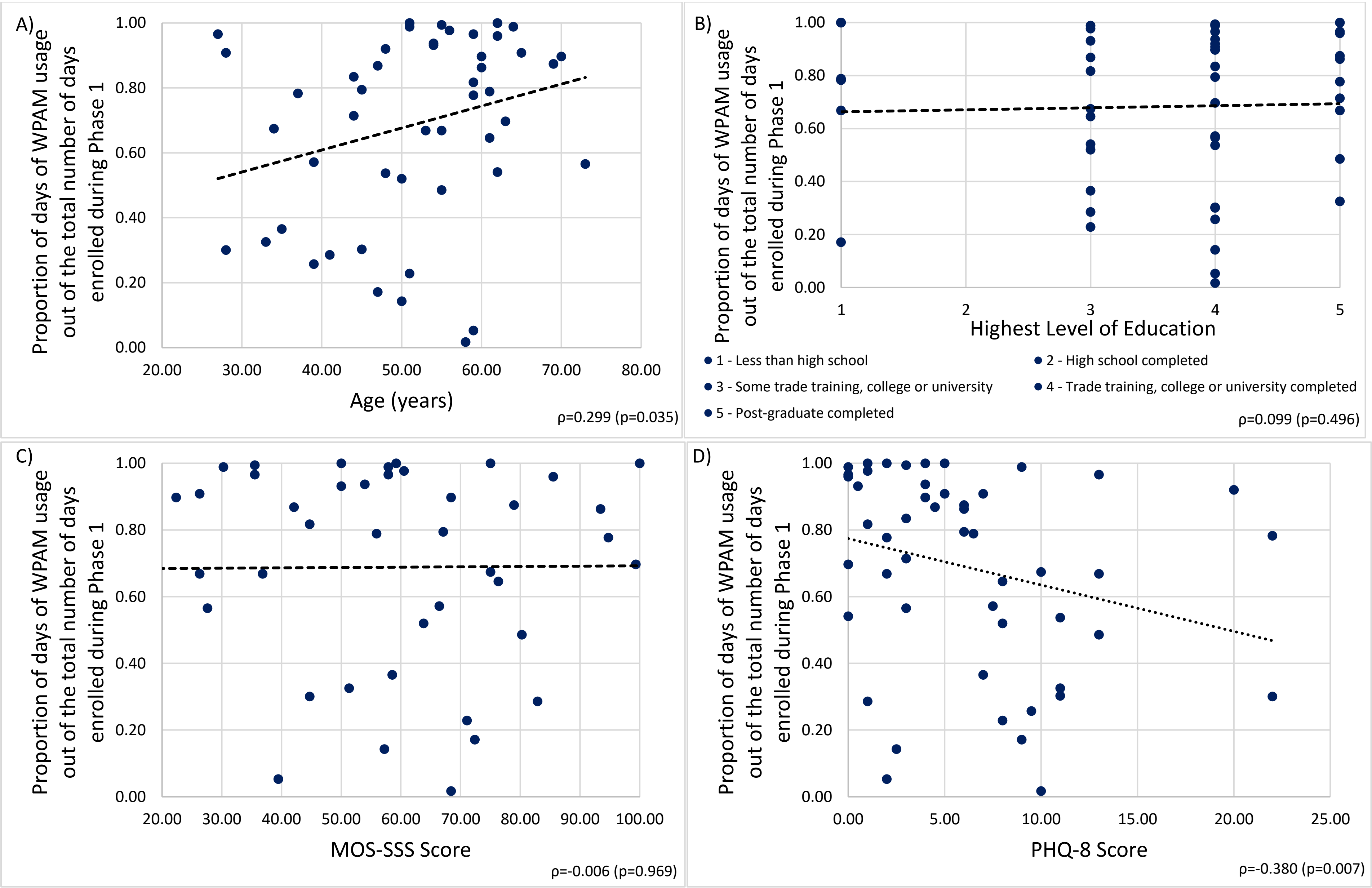


**Supplemental File 3–**In Phase 1 and A) age (n=50), B) highest level of education (n=50), C) social support as assessed by the Medical Outcomes Survey-Social Support Scale (MOS-SSS) (n=49), and D) mental health concerns as assessed by the eight-item Patient Health Questionnaire (PHQ-8) (n=49).

Date Last Revised: September 25, 2022
