## Supplemental File 4 for "Wireless physical activity monitor use among adults living with HIV in a community-based exercise intervention study: a quantitative longitudinal observational study"

### *Supplemental File 4 – Research Ethics Board – University of Toronto*
### *Human Participant Ethics Protocol*

#### 0 - Identification

**RIS Human Protocol Number**
**41886**

**Protocol Title**
MScPT Fitbit Study - Wireless physical activity monitor use among adults living with HIV in a community-based exercise intervention study

**Protocol Type**
**Investigator Submission**

##### Applicant Information

**Applicant Name**
**Miss Kelly O'Brien**

**Rank / Position**
Assoc Professor

**Department / Faculty**
Dept of Physical Therapy - Temerty Faculty of Medi

**Business Telephone**
416-978-0565

**Extension**

**Email Address**


##### Collaborators/Co-Investigators

| Name | Department | Designation |
|---|---|---|
| CARUSONE , SOO CHAN | CASEY HOUSE | Co-Investigator |
| Michael Sperduti | Physical Therapy - MScPT Scholarly Practice Curriculum | Student |
| Josh Turner | Physical Therapy - MScPT Scholarly Practice Curriculum | Student |
| Hayley Sevigny | Physical Therapy - MScPT Scholarly Practice Curriculum | Student |
| Farhanna Hassanali | Physical Therapy - MScPT Scholarly Practice Curriculum | Student |
| Justin Cheng | Physical Therapy - MScPT Scholarly Practice Curriculum | Student |
| Judy Chow | Physical Therapy - MScPT Scholarly Practice Curriculum | Student |
| Matthieu Dagenais | Brock University | Co-Investigator |
| Kelly O’Brien | Dept of Physical Therapy | Principal Investigator |

##### Projected Project Dates

Estimated Start Date
1-Dec-21

Estimated End Date
31-Dec-22



**OFFICE OF RESEARCH ETHICS**
McMurrich Building, 12 Queen’s Park Crescent West, 2nd Floor, Toronto, ON M5S 1S8 Canada
Tel: +1 416 946-3273 ● Fax: +1 416 946-5763 ● ● http://www.research.utoronto.ca/for-researchers-administrators/ethics

#### 2 - Location

**Location of the Research:** ☒ University of Toronto ☐ Other Locations

##### Administrative Approval/Consent

Administrative Approval/Consent Needed: ○ Yes ◉ No

Community Based Particatory Research Project? ○ Yes ◉ No

##### Other Ethic Boards Approval(s)

Another Institution or Site involved? ○ Yes ◉ No

#### 3 - Agreements and Reviews

##### Funding

Project Funded? ◉ Yes ○ No

##### External Funds Administered by U of T

| App No. | Fund No. | Sponsor/Program | Status | Fund End Date | Peer Reviewed |
|---|---|---|---|---|---|
| 179109 | 503961 | CRC - CIHR | Awarded | 2022-12-31 | X |

##### Agreements

Funding/non-funding Agreement in Place? ○ Yes ◉ No

Any Team Member Declared Conflict of Interest? ○ Yes ◉ No

##### Reviews

☒ This research has gone under scholarly review by thesis committee, departmental review committee, peer review committee, or some other equivalent

Type of Review : -e.g.: departmental research committee, supervisor, CIHR, SSHRC, OHTN, etc.

This study protocol was reviewed by research advisors in the MScPT research curriculum, University of Toronto. This study protocol involves using data co

☒ This review was specific to this protocol

☒ The review was part of a larger grant

☐ This research will go under scholarly review prior to funding

☐ This review will not go under a scholarly review

#### 4 - Potential Conflicts

##### Conflict of Interest

Will researchers, research team members, or immediate family members receive any personal benefit? ○ Yes ◉ No

##### Restrictions on Information

Are there any restrictions regarding access to, or disclosure of information (during or after closure)? ○ Yes ◉ No

##### Researcher Relationships


**OFFICE OF RESEARCH ETHICS**
McMurrich Building, 12 Queen's Park Crescent West, 2nd Floor, Toronto, ON M5S 1S8 Canada
Tel: +1 416 946-3273 ● Fax: +1 416 946-5763 ● ● http://www.research.utoronto.ca/for-researchers-administrators/ethics

Are there any pre-existing relationships between the researchers and the researched? ◯ Yes ◉ No

**Collaborative Decision Making**

Is this a community based project - i.e.: a collaboration between the university and a community group? ◯ Yes ◉ No

#### 5 - Project Details

**Summary**

**Rationale**

Describe the purpose and scholarly rationale for the project

RATIONALE:
While physical activity can improve health outcomes in people living with HIV,1,2 studies have shown a wide range of physical inactivity levels in people living with HIV spanning from 19-73%3 and sedentary time of approximately 9 hours a day.4 Community based exercise (CBE) programs can help to increase physical activity and promote well-being in people living with HIV.5 However, strategies are needed to encourage sustained, long-term engagement in physical activity.5 Wireless physical activity monitors (WPAMs) can help to promote and objectively measure physical activity in the general population.6 Evidence on WPAM use in the context of HIV has primarily focused on WPAM as a measurement tool of physical activity.7 To our knowledge, a paucity of research exists that examines the extent of WPAM use and factors that may influence their use among people living with HIV. An understanding of WPAM use among people living with HIV will help to inform their potential to measure and promote physical activity in this population.
PURPOSE:
The purpose of this study is to examine WPAM use among adults living with HIV engaged in a CBE intervention.
OBJECTIVES:
Primary Objectives:
1) To examine the extent of WPAM use (uptake and usage) among adults living with HIV engaged in a 25-week CBE intervention and 32 weeks following a CBE intervention.
2) To examine associations between age, highest level of education, social support, and self-reported mental health with WPAM usage among people living with HIV engaged in a 25-week CBE intervention.
Secondary objectives:
3) To describe physical activity levels as measured by a WPAM (number of steps taken per day) and self-reported aerobic physical activity (physical activity questionnaire), among adults living with HIV engaged in a 25-week CBE intervention.
4) To examine the association between a self-reported measure of aerobic physical activity (physical activity questionnaire) and an objective measure of physical activity (WPAM: number of steps taken) among adults living with HIV engaged in a 25-week CBE intervention.
HYPOTHESES:
Objective 2 - We hypothesize that older age, higher level of education, higher social support, and better self-perceived mental health will each demonstrate a positive moderate association ($0.4 \leq r \leq 0.59$)8 with greater WPAM usage among adults living with HIV during a 25-week CBE intervention.
Rationale: Factors of WPAM use in general populations have demonstrated weak correlations with age, level of education, self-reported health and social support.9,10 Despite the weak associations, people living with HIV may be living with a higher prevalence of mental health conditions11,12 and lower levels of education.13 Also, the majority of the sample are older adults (≥50 years of age) living with HIV, and may have less social support than their younger counterparts.14 Thus, we expect moderate associations between these factors and WPAM use among people living with HIV.
Objective 4 - We hypothesize that self-reported physical activity will demonstrate a positive moderate association ($0.4 \leq r \leq 0.59$)8 with greater WPAM-measured physical activity among adults living with HIV engaged in a 25-week CBE intervention.
Rationale: Authors reported moderate correlations between self-reported physical activity (IPAQ) and total activity counts measured by the actigraph in people living with HIV,15 and a strong correlation between the IPAQ and steps per day measured by a pedometer in Hispanics living with HIV.16 We estimate our participants will be self-aware of their physical activity due to their enrollment in the CBE intervention resulting in accurate self-report, thus supporting our hypothesis of a moderate correlation.
REFERENCES:

1. O'Brien KK, Tynan AM, Nixon SA, Glazier RH. Effectiveness of aerobic exercise for adults living with HIV: systematic review and meta-analysis using the Cochrane Collaboration protocol. BMC Infect Dis. 2016 Apr 26;16:182. PMCID: PMC4845358
2. Voigt N, Cho H, Schnall R. Supervised Physical Activity and Improved Functional Capacity among Adults Living with HIV: A Systematic Review. J Assoc Nurses AIDS Care. 2018 May 31;29(5):667–680. PMCID: PMC7089845
3. Schuelter-Trevisol F, Wolff FH, Alencastro PR, Grigoletti S, Ikeda ML, Brandão ABM, Barcellos NT, Fuchs SC. Physical activity: do patients infected with HIV practice? How much? A systematic review. Curr HIV Res. 2012 Sep;10(6):487–497. PMID: 22762420
4. Vancampfort D, Mugisha J, De Hert M, Probst M, Stubbs B. Sedentary Behavior in People Living With HIV: A Systematic Review and Meta-Analysis. J Phys Act Health. 2017 Jul;14(7):571–577. PMID: 28290767
5. Montgomery CA, Henning KJ, Kantarzhi SR, Kideckel TB, Yang CFM, O'Brien KK. Experiences participating in a community-based exercise programme from the perspective of people living with HIV: a qualitative study. BMJ Open. 2017 Apr 4;7(4):e015861. PMCID: PMC5387963
6. Brickwood KJ, Watson G, O'Brien J, Williams AD. Consumer-Based Wearable Activity Trackers Increase Physical Activity Participation: Systematic Review and Meta-Analysis. JMIR Mhealth Uhealth. 2019 Apr 12;7(4):e11819. PMCID: PMC6484266
7. Dagenais M, Cheng D, Salbach NM, Brooks D, O'Brien KK. Wireless physical activity monitor use among adults living with HIV. Rehabil Oncol. 2019 Jan;37(1):17–28.
8. Campbell MJ, Swinscow TDV. 11 Correlation and Regression. Statistics at Square One. 11th ed. Chichester, UK: BMJ Books; 2009. p. 192.
9. Lee PH, Macfarlane DJ, Lam TH. Factors associated with participant compliance in studies using accelerometers. Gait Posture. 2013 Sep;38(4):912–917. PMID: 23688408
10. Cato MS, Wyka K, Ferris EB, Evenson KR, Wen F, Dorn JM, Thorpe LE, Huang TT-K. Correlates of accelerometry non-adherence in an economically disadvantaged minority urban adult population. J Sci Med Sport. 2020 Aug;23(8):746–752. PMID: 32085979
11. Kendall CE, Wong J, Taljaard M, Glazier RH, Hogg W, Younger J, Manuel DG. A cross-sectional, population-based study measuring comorbidity among people living with HIV in Ontario. BMC Public Health. 2014 Feb 13;14:161. PMCID: PMC3933292
12. Do AN, Rosenberg ES, Sullivan PS, Beer L, Strine TW, Schulden JD, Fagan JL, Freedman MS, Skarbinski J. Excess burden of depression among HIV-infected persons receiving medical care in the united states: data from the medical monitoring project and the behavioral risk factor surveillance system. PLoS ONE. 2014 Mar 24;9(3):e92842. PMCID: PMC3963963
13. Pellowski JA, Kalichman SC, Matthews KA, Adler N. A pandemic of the poor: social disadvantage and the U.S. HIV epidemic. Am Psychol. 2013




**OFFICE OF RESEARCH ETHICS**
McMurrich Building, 12 Queen's Park Crescent West, 2nd Floor, Toronto, ON M5S 1S8 Canada
Tel: +1 416 946-3273 ● Fax: +1 416 946-5763 ● ● http://www.research.utoronto.ca/for-researchers-administrators/ethics

Jun;68(4):197–209. PMCID: PMC3700367
14. Emlet CA. An examination of the social networks and social isolation in older and younger adults living with HIV/AIDS. Health Soc Work. 2006 Nov;31(4):299–308. PMID: 17176977
15. Fillipas S, Cicuttini F, Holland AE, Cherry CL. The international physical activity questionnaire overestimates moderate and vigorous physical activity in HIV-infected individuals compared with accelerometry. J Assoc Nurses AIDS Care. 2010 Apr;21(2):173–181. PMID: 20116301
16. Ramírez-Marrero FA, Rivera-Brown AM, Nazario CM, Rodríguez-Orengo JF, Smit E, Smith BA. Self-reported physical activity in Hispanic adults living with HIV: comparison with accelerometer and pedometer. J Assoc Nurses AIDS Care. 2008 Aug;19(4):283–294. PMID: 18598903

**Methods**

Describe formal/informal procedures to be used

This study protocol involves using data collected as part of the larger community-based exercise (CBE) study (REB protocol approved: # 32910). This study does not require any recruitment of participants nor primary data collection. All data required to address the study objectives are included in the larger CBE study dataset.
STUDY DESIGN:
We will conduct a quantitative longitudinal observational study using data collected from the Community-Based Exercise (CBE) study, that examined the implementation of a CBE intervention with adults living with HIV in Toronto, Ontario (REB Protocol #: 32910). The CBE study included three phases over 22 months: a baseline monitoring phase (Phase 0) which served as the 'control' phase of the study (8 months); a CBE intervention (Phase 1) that included thrice weekly exercise, supervised weekly by a fitness trainer, and monthly self-management education sessions (6 months); followed by follow-up monitoring (Phase 2) whereby participants were expected to continue exercising thrice weekly independently (8 months). The wireless physical activity monitor (WPAM) was introduced at Phase 1 initiation, whereby each participant was provided a Fitbit Zip and asked to wear it daily to measure and provide feedback on their physical activity. As a result, the focus of our study will be Phase 1 (intervention) and Phase 2 (follow-up) of the larger CBE study. See Appendix A for an overview of the CBE study timeline, points of previous data collection, and data sources as they pertain to our study.
Community Based Exercise Intervention:
Phase 1: CBE Intervention Phase (25 weeks):
Participants were provided a YMCA membership and expected to exercise thrice weekly, including a combination of aerobic, resistive, neuromotor and flexibility training for 90 minutes each, supervised weekly by a personal fitness instructor at the Central Toronto YMCA. The type, frequency and duration of CBE sessions were adapted based on participant's goals, interests and abilities. Participants were asked to attend monthly group educational sessions on topics from a health professional or a leader in the HIV community. See the CBE study protocol for further details.
Fitbit Zip:
Participants were offered a Fitbit Zip, clip-on WPAM at Phase 1 initiation. The Fitbit Zip reports daily step count (number of steps taken), walking distance (km), calories burned (calories), and active minutes (minutes spent engaging in an activity that burns at least three metabolic equivalents for a minimum period of 10 minutes). If participants consented to the Fitbit component of the intervention, they were asked to wear the Fitbit Zip on their waist all day except when sleeping, showering, bathing, or swimming, to monitor physical activity levels throughout the week (Appendix B – Fitbit Participant Protocol – that was approved by original CBE Study REB Protocol # 32910). The research team set up a Fitbit account for each participant who was instructed to synchronize their Fitbit Zip to their account (either using a computer or smartphone) weekly. If the Fitbit Zip was lost during the study, it was not replaced. If participants had their own personal WPAM prior to enrollment in the study, they were given the preference to use their own WPAM or the Fitbit Zip for the study. At the end of the study the participants were able to keep their Fitbit Zip.
Phase 2: Follow-up Monitoring Phase (32 weeks): Participants received an 8-month extension to their YMCA membership and were encouraged to continue to engage in unsupervised physical activity thrice weekly without the weekly personal supervision of fitness sessions. Participants were asked to continue to wear and synchronize their Fitbit Zip weekly.
DATA SOURCES:
In this study, will use data from the CBE study from sources such as the synchronized Fitbit Zip data (any daily data tracked such as step count, distance walked, calories burned, and active minutes) and self-reported questionnaires that include: a baseline demographic questionnaire (age, gender, living situation, highest level of education, income, employment status, ethnocultural background, year of HIV diagnosis, viral load, anti-retroviral medication history, HIV care, smoking history), after-baseline demographic questionnaire (number and type of concurrent health conditions, general health status, exercise history and status), weekly exercise logs (participant self-reported Fitbit Zip/WPAM use or non-use to track physical activity), the Medical Outcomes Study-Social Support Scale (MOS-SSS; social support), the Patient Health Questionnaire (PHQ-8; mental health), and the Rapid Assessment of Physical Activity questionnaire (RAPA; self-reported aerobic physical activity). See Appendix A for a timeline of data collection sources that will be used from the larger CBE data set. See Appendix C for our study concepts, definitions and how they will be operationalized (measured) in this study.
Concepts and Measurement:
We will define WPAM use as the uptake and usage of the Fitbit Zip among adults living with HIV engaged in the CBE study.
Uptake: We will define Fitbit Zip uptake as i) the number (%) of participants who agreed to use a Fitbit Zip to monitor their physical activity at Phase 1 initiation of the intervention, ii) the number (%) of participants who were using the Fitbit Zip at Phase 1 completion and agreed to continue to use the Fitbit at Phase 2 initiation, and iii) the number (%) of participants who were using the Fitbit Zip at study completion (out of the total number of participants enrolled in the study). We will report participant reasoning for Fitbit Zip withdrawal (refusal or cease) from use (Appendix C).
Usage: We will define Fitbit Zip usage as the percentage of days each participant wore the Fitbit Zip during the total number of days enrolled in the CBE study. We will measure usage by calculating the number of days each participant (who agreed to use the Fitbit Zip) wore the Fitbit Zip (any daily data tracked such as step count, distance walked, calories burned, or active minutes) divided by the total number of days the participant was enrolled in the study. Furthermore, we will report on the participant's self-reported weekly use or non-use and synchronization of the Fitbit Zip or other WPAM as captured in the Weekly Exercise Logs during the CBE Intervention (Phase 1) (Appendix C).
Contextual Factors of Fitbit Zip Use (Objective 2)
We will examine associations between Fitbit Zip usage and age, level of education, self-reported mental health, and social support among adults living with HIV during the 25-week CBE intervention (Phase 1) (Appendix A).
Age: We will define age as the participant's self-reported age, in years, at CBE study enrollment, as measured by the Baseline Demographic Questionnaire at enrollment of the larger CBE study. (Appendix C).
Highest Level of Education: We will define level of education as the highest level of education achieved by each participant at study enrollment as measured by the Baseline Demographic Questionnaire (at enrollment) with the following options: no formal education, less than grade 9, completed high school, completed trade or technical training, completed college, completed university, or postgraduate education (Appendix C). Age and level of education will be measured at study enrollment as this is the only time these questions were asked in the CBE study.
Social Support: We will define social support as a person's connectedness to one or multiple people who offer physical, emotional and relational support. We will measure social support using the MOS-SSS, a 19-item questionnaire developed to measure self-reported social support in healthy adults. Each item asks the user to rate the frequency of a situation on a 5-point scale ranging from "none of the time" worth 1 point to "all of the time" worth 5 points. Scores range from 19 to 95 with higher scores indicating a higher level of social support. The MOS-SSS was administered bimonthly during the CBE study. We will measure social support using participant's median MOS-SSS score of all their documented scores throughout Phase 1 (months 0, 2, 4, and 6) in the CBE dataset to account for missing data and fluctuations in the participants' social support during the intervention (Appendix C).
Mental Health: We will define mental health as a person's emotional, psychological and social well being. We will measure mental health using the PHQ-8, an 8-


OFFICE OF RESEARCH ETHICS
McMurrich Building, 12 Queen's Park Crescent West, 2nd Floor, Toronto, ON M5S 1S8 Canada
Tel: +1 416 946-3273 ● Fax: +1 416 946-5763 ● ● http://www.research.utoronto.ca/for-researchers-administrators/ethics

item questionnaire developed to assess depressive symptoms in the general population. The PHQ-8 asks users to report to what degree a problem applies to them on a 4-point scale, ranging from “not at all”, worth 0 points to “nearly every day” worth 3 points. Total scores range from 0-24, with higher scores indicating poorer mental health. The PHQ-8 has been used in people living with HIV and was administered bimonthly during the CBE study. We will measure mental health using participant’s median PHQ-8 score of all their documented scores throughout Phase 1 (months 0, 2, 4, and 6) in the CBE study dataset to account for missing data and fluctuations in the participants' mental health during the intervention (Appendix C).
Physical Activity (Objective 3 and 4)
We will measure physical activity defined as objective and self-reported aerobic physical activity among participants in the CBE study. We will measure physical activity using an objective measure (Fitbit Zip) and a self-reported questionnaire (RAPA questionnaire).
Objective Measure of Physical Activity: We will measure physical activity as the daily step count as recorded by the Fitbit Zip. In people living with HIV, the Fitbit Zip demonstrated excellent validity (ICC=.99; IQR of .98-.99) for step count regardless of walking speed, but poor validity (ICC=.20; IQR of −.08 to .47) for distance travelled, when compared to gold-standard measures. Thus, we will use daily step count as measured by the Fitbit Zip to objectively measure physical activity due to its validity in people living with HIV. We will measure daily step count using Fitbit data captured during Phase 1 and 2 of the CBE study (Appendix C).
Self-Reported Measure of Physical Activity: We will also measure physical activity using the RAPA. The RAPA is a 9-item questionnaire divided into two sections: the RAPA-1, which measures aerobic physical activity and the RAPA-2, which measures strength and flexibility. Since the Fitbit Zip measures aerobic physical activity, we will use the RAPA-1 to measure physical activity for our study. The RAPA-1 contains 7 items, asking about the level of engagement in aerobic physical activity. RAPA-1 scores can be divided into five categories; “sedentary” for 1 point, “under-active” for 2 points, “under-active regular – light activities" for 3 points, “under-active regular” for 4-5 points, and “active” for 6-7 points. Higher RAPA-1 scores indicate greater levels of aerobic physical activity. The RAPA has been validated in older adults and used in people living with HIV as a measure of physical activity. The RAPA-1 was administered bimonthly during the CBE study. We will measure physical activity using all documented RAPA-1 scores throughout Phase 1 (months 0, 2, 4, and 6) captured in the CBE study dataset as participants’ physical activity levels may have fluctuated during Phase 1 as a result of the episodic disability experienced by people living with HIV (Appendix C).
DATA ANALYSIS:
We will report participant retention in the CBE study, specifically the number of participants who initiated the CBE intervention, completed the intervention, initiated the follow-up monitoring phase and completed the study. We will report the number of participants who withdrew and reasons for withdrawal (if known). We will use Microsoft Excel and SPSS statistical software to facilitate the analysis. See Appendix D for our data analysis plan.
Participant Characteristics: We will report the demographic characteristics of age, year of HIV diagnosis, viral load (ratio scale) and number of concurrent health conditions (ratio scale) as a median (25-75th percentile). We will describe gender, living situation, source of income, employment status, types of health conditions, ethnocultural backgrounds, HIV care, anti-retroviral medication use (nominal scale) and level of education, general health status, exercise history, exercise status, and smoking history (ordinal scale) as frequencies and percentages.
Distribution of Variables of Interest: We will assess the distribution of the study variables of interest using the Shapiro-Wilk test of normality. This will help to determine whether the data are normally or non-normally distributed to inform the correlational analysis for objective 2.
Objective 1:
Uptake: We will calculate the percentage of Fitbit Zip uptake as the number of participants who agreed to use the Fitbit Zip out of the total number of participants enrolled at: the CBE initiation (month 0), completion of the CBE intervention (month 6), and at end of Phase 2 (month 14). We will report the number (%) of participants using the Fitbit Zip at each of the three time points. We will also report the documented reasons for refusal to use or ceasing use of the Fitbit Zip when documented in the CBE study database.
Usage: We will calculate the Fitbit Zip usage as the number of days each participant used the Fitbit Zip out of the total number of days enrolled in the study. We will calculate Fitbit Zip usage over three time periods in the study: Phase 1, Phase 2, Phase 1 and Phase 2 and calculate the median % (25-75th percentile) of days of Fitbit Zip use across all participants in the study. Furthermore, we will calculate the frequencies and percentages of self-reported weekly use or non-use and synchronization of the Fitbit Zip or other WPAM across the participants that completed Weekly Exercise Logs during the CBE Intervention (Phase 1).
Objective 2: We will calculate four correlation coefficients and their 95% confidence intervals to assess associations between the percentage of days of Fitbit Zip usage (# of days used / total # of days enrolled in the study) during Phase 1, and a) age in years (ratio scale), b) highest level of education (ordinal scale), c) MOS-SSS score (ratio scale), and c) PHQ-8 score (ratio scale). We will calculate a Pearson correlation coefficient for normally distributed ratio variables, and Spearman rho correlation coefficients for ordinal or non-normally distributed ratio variables. We will interpret correlation coefficients as very weak ($r \leq 0.19$), weak ($0.2 \leq r \leq 0.39$), moderate ($0.4 \leq r \leq 0.59$), strong ($0.6 \leq r \leq 0.79$), or very strong ($r \geq 0.8$) with significance of $\alpha$=0.05.
Objective 3: Daily Step Count: For each participant we will calculate their median daily step count (ratio scale) recorded by the Fitbit Zip across all daily step counts recorded during Phase 1. We will calculate the median step daily count (25-75th percentile) of the entire sample during Phase 1.
RAPA-1 Score: For each participant, we will calculate the median RAPA-1 score across all measures recorded during Phase 1 (month 0, 2, 4, and 6 scores). We will calculate the median RAPA-1 score (25-75th percentile) of the entire sample during Phase 1.
Objective 4: We will calculate correlation coefficients and their 95% confidence interval to assess the association between the median daily step count as measured by the Fitbit Zip (ratio scale) and the median RAPA-1 score (ratio scale) across all data points recorded in Phase 1 (month 0, 2, 4, and 6). We will calculate a Pearson correlation coefficient (if normally distributed step count and RAPA scores) or a Spearman rho correlation coefficient (if non-normally distributed). We will interpret correlation coefficients as very weak ($r \leq 0.19$), weak ($0.2 \leq r \leq 0.39$), moderate ($0.4 \leq r \leq 0.59$), strong ($0.6 \leq r \leq 0.79$), or very strong ($r \geq 0.8$) with significance of $\alpha$=0.05.
APPENDICES
Appendix A – Timeline of CBE Intervention and Data Collection for the Community-Based Exercise Intervention Fitbit Study – Date Last Revised: November 1, 2021
Appendix B – Fitbit Participant Protocol – Date Last Revised: November 1, 2021
Appendix C – Definition of Concepts and Demographic Characteristics of Interest in the CBE Fitbit Study – Date Last Revised: November 1, 2021
Appendix D –Data Analysis Plan – Date Last Revised: November 1, 2021
Appendix E – Community-Based Exercise Study Participant Flow Chart – Date Last Revised: November 1, 2021
REVISION - NOVEMBER 26, 2021
APPENDIX BB - Information Letter and Cosent Form from original CBE Study (Approved) - Date Last Revised: March 3, 2017

Copies of questionnaires, interview guided and/or other instruments used

| Document Title | Document Date |
| --- | --- |
| Not Applicable | |

**Clinical Trials**

Is this a clinical trial? ◯ Yes ◉ No


**OFFICE OF RESEARCH ETHICS**
McMurrich Building, 12 Queen’s Park Crescent West, 2nd Floor, Toronto, ON M5S 1S8 Canada
Tel: +1 416 946-3273 ● Fax: +1 416 946-5763 ● ● http://www.research.utoronto.ca/for-researchers-administrators/ethics

#### 6 - Participants and Data

Participants and/or Data

What is the anticipated sample size of number of participants in the study? 69

Describe the participants to be recruited, or the individuals about whom personally identifiable information will be collected. List the inclusion and exclusion criteria. Where the research involves extraction or collection personally identifiable information, please describe where the information will be obtained, what it will include, and how permission to access said information is being sought.

This study involves using data collected from the Community Based Exercise (CBE) conducted by O'Brien et al. (Uof T approved protocol REB # 32910). Data from the CBE study is stored on a secured server at the Episodic Disability and Rehabilitation Research Lab at the University of Toronto. Access to the data will be granted by the lead faculty advisor, Kelly O'Brien. The student research team will use an anonymized version of the data set in order to address the study objectives.
INCLUSION CRITERIA: Participants in this study include adults living with HIV (18 years or older) who enrolled in the CBE study and initiated the Phase 1 CBE intervention.
RECRUITMENT: There is no recruitment of participants required for this study as it involves using data already collected as part of the larger CBE study.
SAMPLE SIZE:
Eighty participants initiated Phase 1, and met the eligibility criteria for inclusion in our study. Of the 80 participants, 67 participants completed Phase 1, and 52 participants completed Phase 2. See Appendix E for the CBE study participant flow chart. Of the 80 participants who initiated the CBE intervention (Phase 1), 69 consented to Fitbit uptake (2021 email from KK O'Brien to us; unreferenced). Thus, our maximum sample size is 69 participants for this study.
We used our primary analytical objective to determine our level of power in which to detect an association between Fitbit use and the contextual factors (objective 2). Using a statistical significance at $\alpha$=0.05 and $\beta$=0.1, a sample size of 69 participants will allow for us to detect a moderate correlation of 0.4 or greater.

Is there any group or individual-level vulnerability related to the research that needs to be mitigated (for example, difficulty understanding consent, history of exploitation by researchers, or power differential between the researcher and the potential participant)? ○ Yes ◉ No

**Recruitment**

Is there recruitment of participant? ○ Yes ◉ No

Is participant observation used? ○ Yes ◉ No

Will translation materials be used/required? ○ Yes ◉ No

Attach copies of all recruitment posters, flyers, letters, email text, or telephone scripts

| Document Title | Document Date |
|---|---|
| Not Applicable | |

Compensation

Will the participants receive compensation? ○ Yes ◉ No

Non Compensation Description

This study pertains to analysis using data from the larger CBE study (REB protocol #32910). There is no recruitment of new participants or collection of new data

Is there a withdrawal clause in the research procedure? ○ Yes ◉ No

#### 7 - Investigator Experience

**Investigator Experience with this type of research**

Please provide a brief description of the previous experience for this type of research by the applicant, the research team, and any persons who will have direct contact with the applicants. If there is no previous experience, how will the applicant and research team be prepared?

Lead faculty advisor: Kelly O'Brien is a physical therapist and Associate Professor in the Department of Physical Therapy at the University of Toronto. Dr. O'Brien previously completed the CBE study in partnership with the Central YMCA (Protocol #32910) to examine the effect of a CBE intervention with adults living with HIV. This research project will be a sub-study using data from the CBE study.
MScPT Students: This study will be conducted by MScPT students (Justin Cheng, Judy Chow, Farhanna Hassanali, Hayley Sevigny, Michael Sperduti, and Joshua Turner) in the Department of Physical Therapy at the University of Toronto in partial fulfillment of the requirements for an MScPT degree. The MScPT student researchers have undertaken Unit 7: Scholarly Practice I of the MScPT curriculum in September 2021, which involved course work on ethics, research strategies, literature review, study design, and protocol development. Dedicated time on Mondays between January to April 2022 is reserved for the MScPT


OFFICE OF RESEARCH ETHICS
McMurrich Building, 12 Queen's Park Crescent West, 2nd Floor, Toronto, ON M5S 1S8 Canada
Tel: +1 416 946-3273 ● Fax: +1 416 946-5763 ● ● http://www.research.utoronto.ca/for-researchers-administrators/ethics

students to work on and attend class sessions related to the data analysis component of the research project. MScPT students will prepare by reviewing relevant statistics concepts and learning to use statistics software such as SPSS, to perform the statistical analyses required for this project. The MScPT students will undertake Unit 13: Scholarly Practice II of the MScPT curriculum, which will involve writing the manuscript and presentation of the research findings on Research and Awards Day in the Department of Physical Therapy at the University of Toronto. A research plan and timeline has been established by the MScPT student researchers to facilitate completion of this research project. Research advisors overseeing this project will provide assistance as needed.
Co-advisor: Soo Chan Carusone is currently affiliated with McMaster University and was the previous Research Lead at Casey House in Complex Lives and HIV research. Dr. Chan Carusone is one of the lead researchers involved in the primary CBE study, and has experience advising on multiple past MScPT student research projects with Kelly O'Brien. Soo is an advisor for this project.
Co-advisor: Matthieu Dagenais is a PhD candidate at Brock University and past MSc graduate in the Rehabilitation Sciences Institute at the UofT (Supervisor: Kelly O'Brien). Matthieu's MSc thesis involved assessing the validity of the Fitbit Zip to measure physical activity in adults living with HIV as part of the CBE study. Matthieu also completed a scoping review on WPAM use among adults living with HIV. Matthieu Dagenais is an advisor for this project and will assist with statistical analyses using SPSS.

Are community members collecting and/or analyzing data? ○ Yes ◉ No

#### 8 - Possible Risks and Benefits

##### Possible Risks

Potential Risk Details:

| | | |
|---|---|---|
| Physical Risks | ○ Yes | ◉ No |
| Psychological/emotional Risks | ○ Yes | ◉ No |
| Social Risk | ◉ Yes | ○ No |
| Legal Risk | ○ Yes | ◉ No |

Risk Description

As data for this research project is stored electronically, there is a risk of loss of privacy if the data is not treated with confidentiality or are not stored and/or transferred securely. In order to mitigate these risks, personal identifiers from the collected data were removed from the CBE study data set and participants were assigned a numeric code to anonymize their data during the CBE study (REB Protocol #32910). This data is stored in a password protected folder on a secured server in the Episodic Disability and Rehabilitation Research Lab at the University of Toronto. Only the MScPT students and the research advisors on this application will have access to this secured server in person on the computers in the Episodic Disability and Rehabilitation Research Lab or remotely using a secured virtual private network (VPN) to access the secured server. Any files related to this study involving data will not be copied or downloaded onto personal computers.
The document that links the participant's numeric code to their name is stored in a password protected folder on the University of Toronto secured server. The student research team will not have access to the personal information of participants, with the exception of the names and emails of participants who agreed to be contacted with a summary of the study findings. Kelly O'Brien will provide this information at the end-of-grant KTE of the study.
All hard-copy documents from the CBE study, including consent forms and questionnaires that were completed throughout the study by participants, are stored in a locked filing cabinet in the Episodic Disability and Rehabilitation Research Lab at the University of Toronto.

##### Potential Benefits

Benefit Description

While there are no direct benefits to participants in the study, results may help to improve our understanding of WPAM use in a community-dwelling sample of adults living with HIV who engaged in a CBE intervention. A better understanding of WPAM use among people living with HIV will help to inform their potential

#### 9 - Consent

Consent Process Details

All of the participants provided written informed consent to participate in the original CBE study (Protocol #32910), which includes data collected that will be used for this study analysis. Furthermore, participants who consented to receive knowledge translation will receive a summary of the findings through email or other specified contact method. As this study involves analysis of data as part of an existing data set from data collected as part of the original CBE study, we will not seek informed consent as the data required for this study has already collected as part of the CBE study (REB Protocol #32910).

Uploaded letter/consent form(s)

| Document Title | Document Date |
|---|---|
| Appendix BB - Consent Form from Original CBE Study (March 2017) - Protocol - 32910 | 2017-03-03 |

Is there additional documentation regarding consent such as screening materials, introductory letters etc.: ○ Yes ◉ No

Uploaded letter/consent form(s)


**OFFICE OF RESEARCH ETHICS**
McMurrich Building, 12 Queen's Park Crescent West, 2nd Floor, Toronto, ON M5S 1S8 Canada
Tel: +1 416 946-3273 ● Fax: +1 416 946-5763 ● ● http://www.research.utoronto.ca/for-researchers-administrators/ethics

Will any information collected in the screening process - prior to full informed consent to participate in the study - be retained for those who are later excluded or refuse to participate in the study? ( ) Yes (•) No

Is the research taking place within a community or organization which requires formal consent be sought prior to the involvement of the individual participants ( ) Yes (•) No

Are any participants not capable (e.g.: children) of giving competent consent? ( ) Yes (•) No

#### 10 - Debriefing and Dissemination

##### DeBrief

Will deception or intentional non disclosure be used? ( ) Yes (•) No

Will a written debrief be used? ( ) Yes (•) No

Do participants/communities have the right to withdraw their data following the debrief? ( ) Yes (•) No

Information Feed Back Details following completion of a participants participation in the project

We will submit a manuscript to the Department of Physical Therapy as required by the MScPT research curriculum and present research findings at the Department of Physical Therapy Research Day, and other relevant conferences (e.g. Ontario Physiotherapy Association conference) and community organizations (e.g. Casey House). We will submit our manuscript to a peer-reviewed research journal for publication and develop a summary sheet of study findings that we will share via email to study participants who agreed to be informed about the CBE study results.

Procedural details which allow participants to withdraw from the project

[X] Not Applicable

What happens to a participants data and any known consequences related to the removal of said participant

[X] Not Applicable

List reasons why a participant can not withdraw from the project (either at all or after a certain period of time)

[X] Not Applicable

#### 11 - Confidentiality and Privacy

##### Confidentiality

Is the data confidential? (•) Yes ( ) No

Will the confidentiality of the participants and/or informants be protected? (•) Yes ( ) No

List confidentiality protection procedures

Personal identifiers from the collected data were removed from the CBE study data set and participants were assigned a numeric code to anonymize their data during the CBE study. All electronic files are stored in a password protected folder on a secured server in the Episodic Disability and Rehabilitation Research Lab at the University of Toronto. Only the MScPT students and the research advisors on this application will have access to this secured server in person on the computers in the Episodic Disability and Rehabilitation Research Lab or remotely using a secured virtual private network (VPN) to access the secured server. Any files related to this study involving data will not be copied or downloaded onto personal computers.

The document that links the participant's numeric code to their name is stored in a password protected folder on the University of Toronto secured server. The student research team will not have access to the personal information of participants, with the exception of the names and emails of participants who agreed to be contacted with a summary of the study findings. Kelly O'Brien will provide this information at the end-of-grant KTE of the study.

Are there any limitations on the protection of participant confidentiality? ( ) Yes (•) No

Is participant anonymity/confidentiality not applicable to this research project? ( ) Yes (•) No

##### Data Protection



**OFFICE OF RESEARCH ETHICS**
McMurrich Building, 12 Queen's Park Crescent West, 2nd Floor, Toronto, ON M5S 1S8 Canada
Tel: +1 416 946-3273 ● Fax: +1 416 946-5763 ● ● http://www.research.utoronto.ca/for-researchers-administrators/ethics

Describe how the data (including written records, video/audio recordings, artifacts and questionnaires) will be protected during the conduct of the research and subsequent dissemination of results

We will obtain an anonymized data set from the CBE study from Kelly O'Brien (lead faculty advisor). Data from the CBE study is kept on a secure server in the Episodic Disability and Rehabilitation Research Lab at the University of Toronto. The dataset will remain on the secured server in a password-protected folder accessible only to our team listed on this application. We will be able to access the information on the server remotely using a password-protected virtual private network (VPN). Any files related to this study involving data will not be copied or downloaded onto personal computers.
The document that links the participant's numeric code to their name is stored in a password protected folder on the University of Toronto secured server. The student research team will not have access to the personal information of participants, with the exception of the names and emails of participants who agreed to be contacted with a summary of the study findings. Kelly O'Brien will provide this information at the end-of-grant KTE of the study.
All hard-copy documents from the CBE study, including consent forms and questionnaires that were completed throughout the study by participants, are stored in a locked filing cabinet in the Episodic Disability and Rehabilitation Research Lab at the University of Toronto.

Explain for how long, where and what format (identifiable, de-identified) data will be retained. Provide details of their destruction and/or continued storage. Provide a justification if you intend to store identifiable data for an indefinite length of time. If regulatory requirements for data retention exists, please explain.

Electronic data used for this study will be stored in the Episodic Disability and Rehabilitation Research Lab at the University of Toronto. This data will be retained for 10 years after which this information will be destroyed by the lead investigator of the CBE study, Kelly O'Brien.

Will the data be shared with other researchers or users? ○ Yes ● No

#### 12 - Level of Risk and Research Ethics Board

##### Level of Risk for the Project

Group Vulnerability: Low

Research Risk: Low

Risk Level: 1

##### Explanation/Justification

Explanation/Justification detail for the group vulnerabilty and research risk listed above

There is minimal risk to participants in this study as no further data collection from participants is required for this study. This study involves the use of an anonymized data set of existing data collected as part of the larger study.

##### Research Ethics Board

REB Associated with this project: Health Sciences

#### 13 - Application Documents Summary

##### Uploaded Documents

| Document Title | Document Date |
|---|---|
| REB Response to Reviewer Comments | 2021-11-26 |
| Appendix BB - Consent Form from Original CBE Study (March 2017) - Protocol - 32910 | 2017-03-03 |
| Appendix A - Timeline of CBE Intervention and Data Collection for the Community-Based Exercise Intervention Fitbit Study | 2021-11-01 |
| Appendix B - REB Approved Fitibit Protocol (#32910) | 2021-11-01 |
| Appendix C - Definition of Concepts and Demographic Characteristics of Interest in the CBE Fitbit Study | 2021-11-01 |
| Appendix D - Data Analysis Plan | 2021-11-01 |
| Appendix E - Community-Based Exercise Study Participant Flow Chart | 2021-11-01 |

#### 14 - Applicant Undertaking


OFFICE OF RESEARCH ETHICS
McMurrich Building, 12 Queen's Park Crescent West, 2nd Floor, Toronto, ON M5S 1S8 Canada
Tel: +1 416 946-3273 ● Fax: +1 416 946-5763 ● ● http://www.research.utoronto.ca/for-researchers-administrators/ethics

I confirm that I am aware of, understand, and will comply with all relevant laws governing the collection and use of personal identifiable information is research. I understand that for research involving extraction or collection of personally identifiable information, provincial, federal, and/or international laws may apply and that any apparent mishandling of said personally identifiable information, must be reported to the office of research ethics.

As the Principal Investigator of the project, I confirm that I will ensure that all procedures performed in accordance with all relevant university, provincial, national, and/or international policies and regulations that govern research with human participants. I understand that if there is any significant deviation in the project as originally approved, I must submit an amendment to the Research Ethics Board for approval prior to implementing any change.

☒ I have read and agree to the above conditions


**OFFICE OF RESEARCH ETHICS**
McMurrich Building, 12 Queen's Park Crescent West, 2nd Floor, Toronto, ON M5S 1S8 Canada
Tel: +1 416 946-3273 ● Fax: +1 416 946-5763 ● ● http://www.research.utoronto.ca/for-researchers-administrators/ethics

RIS Protocol Number: 41886

Approval Date: 8-Dec-21

PI Name: Kelly O'Brien

Division Name:

Dear Miss Kelly O'Brien:

Re: Your research protocol application entitled, "MScPT Fitbit Study - Wireless physical activity monitor use among adults living with HIV in a community-based exercise intervention study"

The Health Sciences REB has conducted a Delegated review of your application and has granted approval to the attached protocol for the period 2021-12-08 to 2022-12-07.

This approval covers the ethical acceptability of the human research activity; please ensure that all other approvals required to conduct your research are obtained prior to commencing the activity.

Please be reminded of the following points:

An **Amendment** must be submitted to the REB for any proposed changes to the approved protocol. The amended protocol must be reviewed and approved by the REB prior to implementation of the changes.

An annual **Renewal** must be submitted for ongoing research. Renewals should be submitted between 15 and 30 days prior to the current expiry date.

A **Protocol Deviation Report** (PDR) should be submitted when there is any departure from the REB-approved ethics review application form that has occurred without prior approval from the REB (e.g., changes to the study procedures, consent process, data protection measures). The submission of this form does not necessarily indicate wrong-doing; however follow-up procedures may be required.

An **Adverse Events Report (AER)** must be submitted when adverse or unanticipated events occur to participants in the course of the research process.

A **Protocol Completion Report** (PCR) is required when research using the protocol has been completed.

If your research is funded by a third party, please contact the assigned Research Funding Officer in Research Services to ensure that your funds are released.

Best wishes for the successful completion of your research.



**OFFICE OF RESEARCH ETHICS**
McMurrich Building, 12 Queen's Park Crescent West, 2nd Floor, Toronto, ON M5S 1S8 Canada
Tel: +1 416 946-3273 ● Fax: +1 416 946-5763 ● ● http://www.research.utoronto.ca/for-researchers-administrators/ethics
