## Supplemental File 5 for "Wireless physical activity monitor use among adults living with HIV in a community-based exercise intervention study: a quantitative longitudinal observational study"

### Supplemental File 5 - Information Letter and Consent Form

**Title of Study:** Translating Exercise into the HIV Community: Evaluating a Community-Based Exercise Intervention to Improve the Health among Adults Living with HIV

**Principal Investigator:**
Kelly O'Brien, PhD, BScPT
Department of Physical Therapy, University of Toronto
500 University Avenue, Room 160, Toronto, Ontario, M5G 1V7


**Co-Investigator(s):**
Patty Solomon, PhD
School of Rehabilitation Science, Faculty of Health Sciences, McMaster University
1400 Main Street West, IAHS 403, Hamilton, Ontario, L8S 1C7


Ahmed Bayoumi, MD, MSc, FRCPC
St. Michael's Hospital, Centre for Research on Inner City Health
30 Bond Street, Toronto, Ontario, M5B 1W8


Aileen Davis, PhD
Toronto Western Hospital Main Pavilion
399 Bathurst Street, 11th Floor Rm 322, Toronto, Ontario M5T 2S8


Ada Tang, PhD, PT
School of Rehabilitation Science, Faculty of Health Sciences, McMaster University
1400 Main Street West, IAHS 437, Hamilton, Ontario, L8S 1C7


Sean Rourke, PhD
St. Michael's Hospital, Centre for Research on Inner City Health
30 Bond Street, Toronto, Ontario, M5B 1W8


**Sponsor: This research was funded by the Canadian Institutes of Health Research (CIHR), HIV/AIDS Community-Based Research Program.**

You are invited to participate in a research study to evaluate a Community-Based Exercise (CBE) intervention to improve health among people living with HIV. This study is being conducted in collaboration with the Central Toronto YMCA. In order to decide whether or not you want to be a part of this research study, you should understand what is involved and the potential risks and benefits. This form gives detailed information about the research study, which will be discussed with you. Once you understand the study, you will be asked to sign this form if you wish to participate. Please take your time to make your decision.

**WHY IS THIS RESEARCH BEING DONE?**

HIV is now considered a chronic illness where more individuals are living longer and aging with the health-related challenges of HIV, comorbidities (co-existing health challenges) and potential side effects of treatment. Exercise is one intervention that can help people living with HIV address their health-related challenges and improve their overall health. It is important for an exercise program to be accessible and practical for people living with HIV to sustain over the long term. Community-Based Exercise (CBE) is an ideal model in which to enhance physical activity and health outcomes among people living with HIV.

**WHAT IS THE PURPOSE OF THIS STUDY?**

The purpose of this research is to evaluate a community-based exercise (CBE) intervention for adults living with HIV within the community. We are specifically interested in learning about the extent adults with HIV participate in a CBE program, the effect of CBE on the health of adults living with HIV, and the adherence to exercise. We are also interested in learning from people living with HIV, fitness instructors and managers about the strengths and challenges of implementing a CBE program for people living with HIV in the community.

**WHO CAN PARTICIPATE IN THIS STUDY?**

You are eligible to take part in this study if you are 18 years of age or older living with HIV in Ontario, and consider yourself medically stable and safe to participate in a Community-Based Exercise program. If you are interested in the study you will be asked to complete a Physical Activity Readiness Questionnaire. Your responses to this questionnaire will help determine your ‘readiness’ to exercise and whether you will need to talk with your physician to confirm your ability to take part in physical activity that is involved with this study. We encourage all interested individuals to discuss their participation in this study with their physician (although written documentation from a physician is not required for participation in the study).

**WHAT IS INVOLVED BY TAKING PART IN THE STUDY?**

If you decide to participate in this study, you will be asked to take part in a 22 month study that is broken up into three phases. In order to take part you must commit to take part in all three phases of the study.

**Phase One (8 months): Baseline Monitoring -** The first eight months (32 weeks) will be a monitoring phase in order to obtain a baseline level of your health. You will be asked to attend the YMCA every two months to complete a series of health assessments and questionnaires. The health assessments will include a fitness test where we you will be asked to run on a treadmill or cycle on a stationary bike to measure your oxygen consumption and rate of exertion. We will also measure your heart rate and blood pressure during this test as well. At the same appointment, you will undergo strength testing of major upper and lower body muscle groups. Some of the measures will include grip strength, vertical jump test, and partial curl ups. We will measure your weight and body composition including body mass index, fat composition, and waist and hip circumference. We will also measure your flexibility of your lower back and hamstrings. This physical assessment will take approximately 1.5 hours.

You will also be asked to fill out a series of questionnaires that include questions about your physical, mental and cognitive health. You will be asked to complete a physical activity and a demographic questionnaire that asks about your age, gender and overall health status. We recently moved these questionnaires into a web-based computer format. The questionnaires will be filled out on a computer at the University of Toronto with the Research Coordinator who will be able to help you if needed. If you do not feel comfortable using the computer, you will have the option to continue to complete the questionnaires in a paper format.

At the end of the baseline monitoring phase, just before the exercise intervention phase, you will also be asked to complete a neurocognitive assessment that consists of nine different measures testing your speed and memory. The measures will be assessed using a web-based online cognitive assessment using the NIH Toolbox ® App with an iPad. Completing the questionnaires and the neurocognitive assessment will take approximately 2 hours. You will be able to take breaks throughout as needed. The Research Coordinator will call or email you (depending on your preference) to remind you of any upcoming health assessment appointments.

**Phase Two (6 months): Exercise Intervention -** In this next phase, you will be asked to attend exercise sessions at the YMCA for approximately 1.5 hours three times per week for 24 weeks. The exercise sessions may include a combination of aerobic, strength, and flexibility training and may be done individually or in a group format. A fitness instructor will meet with you and determine an ideal exercise program specifically individualized for you. The sessions will be supervised weekly by a fitness instructor at the YMCA who will monitor your progress and progress exercise intensity. In addition, you will also be asked to attend educational sessions every month (6 sessions in total) at the YMCA focused on topics related to self-management and healthy lifestyle living with HIV. Topics of the six sessions may include: stress and relaxation, pain management, nutrition, fatigue management, continuing physical activity, smoking cessation, goal setting, confidence and self-management, community services and support, living with HIV, rehabilitation in the context of HIV, sexuality and body image, and wrap up and overall Q&A. During this time, you will continue with the above bimonthly health assessments and questionnaires.

**Phase Three (8 months): Self-Monitored Exercise -** In this next phase you will be encouraged to continue to take part in the exercise sessions three times per week at the YMCA for 8 months (32 weeks). However, this time the exercise will be largely unsupervised. During this time, you will continue with the bimonthly health assessments and questionnaires.

During Phases Two and Three, you will be asked to complete a weekly online exercise log to track the frequency, intensity, time and type of exercise that you do. You will also be provided with a Fitbit™ which is a small wireless activity device to self-monitor steps, distance and calories burned.

You will be asked to sync your Fitbit™ weekly to your phone/home computer and the Fitbit information can be used to help you fill out your weekly exercise log. If you do not have access to a computer you can sync your Fitbit information on the computer at the research lab at the University of Toronto.

The exercise log and information from the Fitbit™ will be submitted monthly to the study investigators to track your activity level throughout the study.

During Phase Two, at the end of each of the monthly educational sessions, you will be invited to take part in a feedback focus group discussion. This is so the researchers can obtain feedback from participants on how the CBE intervention and study is going so far and to suggest ways we might improve on the delivery of the CBE intervention. This feedback session is voluntary. If you attend the education session you are not required to stay for the feedback session. If there are some questions that you do not wish to answer for any reason, you are free to do so, and you may stop at any time. The entire discussion will take about 20-30 minutes. The discussion will be audio recorded and then typed out. The typed report will be stored as a computer file at the University of Toronto and printed onto paper. Your name or other information that could identify who you are will never appear on the audio file, in the computer file or on the printed pages.

**WHAT HAPPENS IF I HAVE AN EPISODE OF ILLNESS?**

Sometimes the episodic nature of HIV may cause situations where your health status may change, which can affect your ability to safely engage in a fitness or questionnaire assessment and affect your ability to continue with the study. If any point during the study, the Research Coordinator or YMCA staff feels that it is not safe for you to start or continue a fitness or questionnaire assessment, they will stop the assessment. In these situations, you will be asked to talk with your physician by phone or in person to discuss your ability to safely engage in physical activity and to continue your participation in the study. You will be asked to report this information to the Research Coordinator who then may reschedule the fitness and/or questionnaire assessments at a time that will be safe and feasible for you.

If you withdraw from the study due to health reasons and would like to re-join the study at a later point in time when your health permits, you will be able to do so if we are still recruiting participants for the study. If we are still in the recruitment phase of the study, the Research Coordinator may schedule a new eligibility and consent meeting with you to ensure you are able to re-join the study. If you re-join the study you will need to start at the baseline monitoring phase again.

If you withdraw from the study due to health reasons and re-join at a later date, the data collected from you prior to your withdrawal will be kept unless you indicate otherwise and request to have your data removed from the study.

**Level of Commitment –** Since this study is 22 months in length, it is important to ensure that participants remain committed to attending the exercise sessions and assessments. We understand the episodic nature of living with HIV and therefore understand last minute cancellations may occur due to health reasons. These circumstances will be taken into consideration when evaluating ongoing eligibility. For this study is it important that we have the assessment information across the 22 months in order to accurately determine the impact of the exercise intervention. So, in order to remain eligible to participate in the study, you will be required to:

- Complete at least three of the four assessments (Fitness and Questionnaire Assessment) during Phase One to continue on to Phase Two. Complete at least three of the four assessments (Fitness and Questionnaire Assessments) during Phase Two to continue on to Phase Three.
- Provide at least 24 hours' notice of cancellation for any Fitness Assessments or exercise sessions with fitness instructors throughout the study. If you do not provide this notice for up to 7 fitness assessments or exercise sessions throughout the 22 month study, you **may** be excluded from the study and your membership will be cancelled (if applicable). This is so that we can recruit additional participants to the study if needed.
- In the event that you have to reschedule a fitness or questionnaire assessment, they must occur within 7 to 10 days of each other. This is to ensure we capture a snapshot of your health (fitness and questionnaire-based) at a point in time.
- Attend exercise sessions three times a week (1.5 hours each) to your best ability.
- Attend the 6 educational sessions in Phase 2 to your best ability.

**WHAT ARE THE POSSIBLE RISKS?**

There is an element of physical risk to participating in this study including injury with exercise and you may perceive the type of questions asked about disability or in the self-reported health questionnaires as 'psychologically probing'. If this does occur, fitness staff from the YMCA will follow appropriate emergency procedures and you will be asked to follow-up with your physician. It is also possible that you may find some of the questions to be personal or sensitive in nature and may also experience loss of privacy during the CBE intervention. You can choose not to complete any of the physical assessments or choose not to answer any questions and you may end the exercise program at any time.

There is a small risk of skin irritation from wearing the Fitbit™, even as instructed on the waistband. If this occurs, we recommend clipping the Fitbit™ on to a belt or pocket, another external form of clothing.

**WHAT ARE THE POSSIBLE BENEFITS?**

Exercise is a beneficial strategy to address health challenges among people living with health conditions as well as enhancing health promotion and disease prevention. Benefits of exercise for people living with HIV include improvements in cardiopulmonary fitness, strength, weight and body composition, and psychological status. CBE programs also have the potential to promote peer support and social interaction in a group context under the supervision of an exercise instructor who can provide ongoing support and encouragement to exercise. Programs may focus on emotional, cognitive and behavioural self-management strategies to help individuals independently manage challenges associated with their chronic condition. This study aims to evaluate these possible benefits of a CBE intervention when implemented with people living with HIV in the community. Lessons learned from this study will be used to guide future implementation of CBE programs for people living with HIV across Canada.

**WHAT IF I DO NOT WANT TO TAKE PART IN THE STUDY?**

You are free to decide if you want to take part in this study or not. If you decide not to take part, you can withdraw at any time. Withdrawing from this study will not affect any of the services that you currently receive at the participating sites in this study. Withdrawing from the study means that the study investigators may still include information collected before you withdrew unless you request for investigators to remove or destroy your information. If you decide to withdraw, we will ask you to complete a study 'exit' and demographic questionnaire, but you can choose not to complete them.

**WHAT INFORMATION WILL BE KEPT PRIVATE?**
All documentation related to the project including completed questionnaires and focus group feedback transcripts will be stored in a locked cabinet at the University of Toronto or on a password protected computer inside a secured office, accessible only to the investigators and research coordinator. Unique identifiers will be used to match questionnaires, Fitbit™ profiles, and cognitive assessments with participants. Only pooled data will be presented in final publications to ensure participant anonymity. It is important to realize that the researchers, research coordinator and fitness instructors and instructors doing the assessments at the YMCA with this study will be aware that you are HIV positive. Given the nature of individual and group exercise or activity, other participants in this study will also know that you are HIV positive. In addition, the Fitness Instructor (Coach) who you are paired with at the Central Toronto YMCA will be provided your Physical Activity Readiness Questionnaire (PARQ), Goal Attainment Scale as well as your phone number and/or e-mail address. The Fitness Instructors (Coaches) will use this information to help inform the program that they develop for you as well as will be in touch with you throughout the intervention regarding scheduling appointments.

The questionnaire information will be administered using Qualtrics Survey Software. This software collects data on a secure server in Canada. The information from the questionnaires will be downloaded from Qualtrics to the secure server at the University of Toronto.

The Fitbit Zip™ and the NIH Toolbox ® App involve collecting data on physical activity and cognitive assessments onto servers in the United States and therefore are subject to U.S. laws, including the U.S. Patriot Act, 2001. As a result, there is a possibility that this information may be accessed by the U.S. government, in compliance with the Patriot Act, without your knowledge or consent. We added a statement in this revised consent form that will enable you to choose whether you wish to opt out the Fitbit™ or Neurocognitive Assessment portions of this study.

As a participant in this research study you will receive a membership to the YMCA. As part of the regular YMCA membership process, the YMCA collects personal and emergency contact information from all of its members (including name, mailing and email address, phone number, birthday, allergies or health concerns, gender, emergency contact name and phone number). This will be collected and stored at the Central Toronto YMCA as part of their general membership process, but is not part of the research study. The YMCA does not provide information about their members to others. However, they do communicate with members regarding YMCA promotions and newsletters. All communication with YMCA members is within the new Canadian Anti-Spam Legislation. Any other information as it relates to the research study will be stored at the University of Toronto.

**WILL I BE PAID TO PARTICIPATE IN THIS STUDY?**
You will not be paid to take part in this study; however as a study participant, you will receive an open access membership to the Central YMCA during the 14 month exercise phase of the study, valued at approximately $1028.02.

The membership will be provided in two waves. The first membership will be for 6 months (intervention phase) and the second membership will be for 8 months (post-intervention phase). To receive the first membership for 6 months, you will need to start the study (complete the baseline assessment), remain in the study for the baseline phase (0-8 months) and complete the bimonthly assessments in the baseline phase. To receive the second membership for 8 months, you will need to remain in the study for the intervention phase (8-14 months) and complete the bimonthly assessments in the intervention phase.

You will also be able to keep the Fitbit™ wireless activity monitor provided in Phase Two as a token of appreciation for your participation in the study.

You will not be paid to take part in the monthly feedback sessions; however there will be food and refreshments available.  If you choose not to take part in the feedback sessions that occur at the end of the educational sessions, you will still receive food and refreshments.

**WILL THERE BE ANY COSTS?**
Your participation in this research project will not involve any additional costs to you other than travel costs (if any) to and from the YMCA.

**WILL THE RESULTS BE PUBLISHED?**
Results of this study will be presented at conferences and published in a scientific journal. We will also develop a fact sheet summary of the results that will be available at participating AIDS Service Organizations (Toronto People with AIDS Foundation), Casey House, the Canadian Working Group on HIV and Rehabilitation and the Central YMCA. The investigators will not include personal information such as your name in the summary so that any publication of results will not identify you. If you are interested in receiving a copy of the study summary, you can contact Kelly O'Brien at.

**IF I HAVE ANY QUESTIONS OR PROBLEMS, WHOM CAN I CALL?**
If you have any questions about the research now or later, please contact Kelly O'Brien (Principal Investigator at the University of Toronto at 416-978-0565. If you have any questions regarding your rights as a research participant, you may contact the Office of the Research Ethics of the University of Toronto at 416-946-3273 or email at.

**CONSENT STATEMENT**
**Translating Exercise into the HIV Community: Evaluating a Community-Based Exercise Intervention to Improve the Health among Adults Living with HIV**

Participant:
I have read the preceding information thoroughly. I have had the opportunity to ask questions, and all of my questions have been answered to my satisfaction. I am committed and agree to participate in this 22 month study. I understand that I will receive a signed copy of this form.

**Fitbit Zip™**

**I DO consent to taking part in the Fitbit™ portion of this study.**
**I DO NOT consent to taking part in the Fitbit™ portion of this study.**

**Cognitive Assessments**

**I DO consent to taking part in the cognitive assessment portion of this study.**
**I DO NOT consent to taking part in the cognitive assessment of this study.**

| **Name of Participant** | **Signature** | **Date** |
|---|---|---|

Person obtaining consent:
I have discussed this study in detail with the participant. I believe the participant understands what is involved in this study.

| Name, Role in Study | Signature | Date |
|---|---|---|

This study has been reviewed by the HIV Research Ethics Board at the University of Toronto. The REB is responsible for ensuring that participants are informed of the risks associated with the research, and that participants are free to decide if participation is right for them. If you have any questions about your rights as a research participant, please call the Office of the Research Ethics of the University of Toronto at **416-946-3273 or email at**
